# Discordant associations of IGF-binding proteins 1 & 2 with diabetes and cardiovascular disease: insights from UK Biobank

**DOI:** 10.64898/2026.07.17.26358347

**Authors:** Emily R Rolfe-Hammerton, Marcella S Conning-Rowland, Lia E De Faveri, Katie J Simmons, Paul J Meakin, Richard M Cubbon, Stephen B Wheatcroft

## Abstract

1.

The insulin-like growth factor (IGF)/IGF-binding protein (IGFBP) axis has been implicated in diabetes mellitus and the associated burden of cardiovascular complications. Higher circulating levels of IGFBP-1 and IGFBP-2 have been established as markers of protection from incident type 2 diabetes, yet their associations with cardiovascular disease remain unclear. Utilising the UK Biobank (UKB) resource to integrate disease outcomes, plasma proteomics and MRI data, we examined associations of IGFBP-1 and IGFBP-2 with incident diabetes and cardiovascular disease. Approximately 50,000 UKB participants with plasma proteomic measurements for IGFBP-1 and IGFBP-2 were included. Multivariate Cox regression models revealed that participants in the highest quartiles of IGFBP-1 and IGFBP-2 had a substantially lower risk of incident diabetes (hazard ratio (HR) = 0.31 and 0.32 respectively), but, paradoxically, had increased risks of incident macrovascular disease, all-cause and cardiovascular-related mortality (HR = 1.81 and 2.39). Both proteins were negatively associated with HbA1c levels, triglyceride/HDL ratio and abdominal adiposity, yet positively associated with NT-proBNP, troponin I, cardiac chamber size and aortic dimensions. In summary, negative associations of IGFBP-1 and IGFBP-2 with incident diabetes mellitus did not translate to a reduced cardiovascular risk, suggesting potentially complex actions of IGFBP-1 and IGFBP-2 in the pathophysiology of cardiometabolic disease.

**Graphical Abstract:** Graphical abstract illustrating the analysis pipeline and aims of this study to identify associations of IGFBP-1 and IGFBP-2 with cardiometabolic disease. Graphical abstract created with BioRender.com.

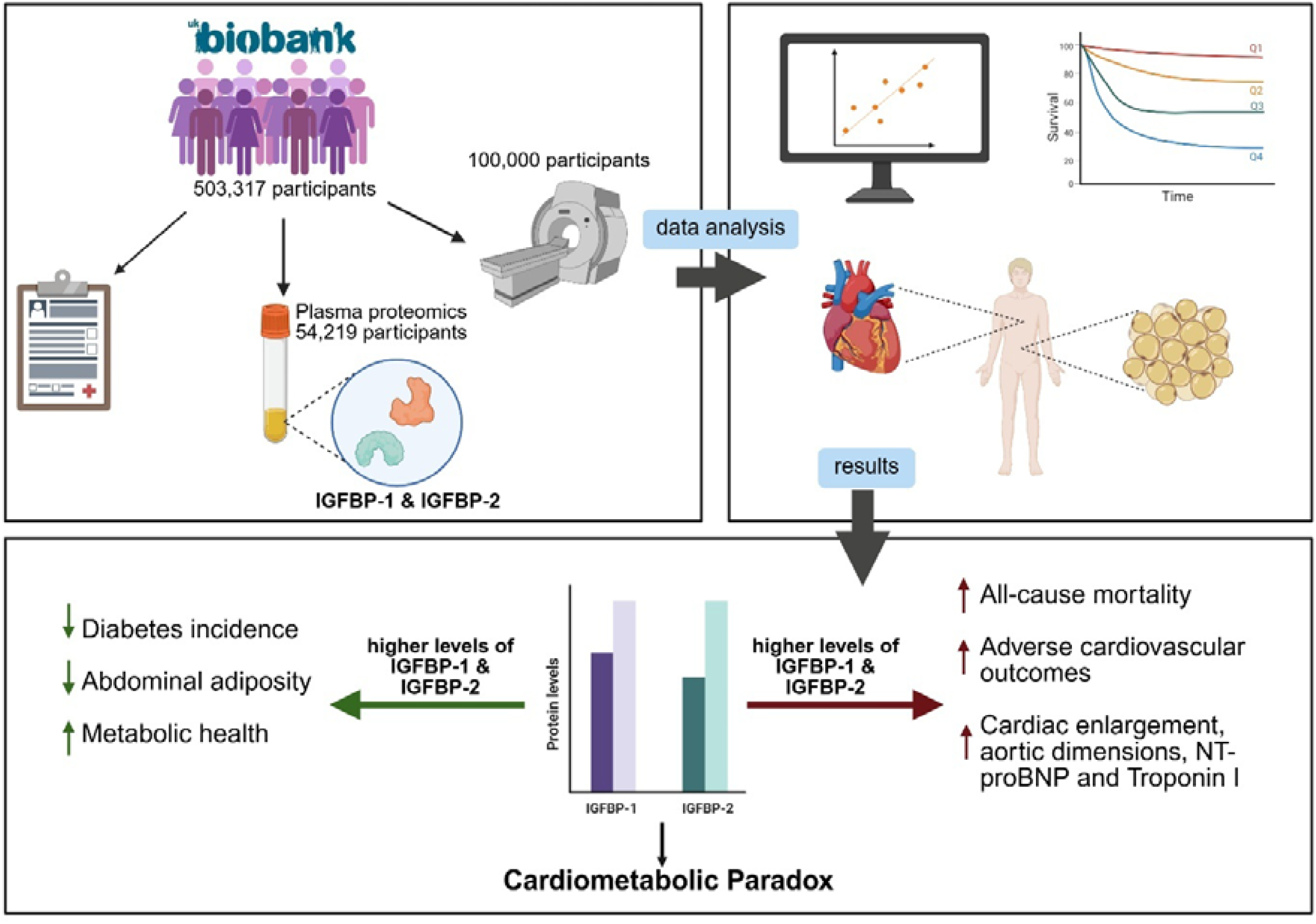

**Article highlights:**

a) The associations of IGFBP-1/-2 with macrovascular disease in cohorts with and without type 2 diabetes are conflicting.
b) Can UK Biobank help to clarify the associations of IGFBP-1 and IGFBP-2 with cardiometabolic disease?
c) Higher levels of both proteins are associated with a reduced risk of diabetes incidence, but an increased risk of cardiovascular disease. This is the first study to integrate survival analyses with large-scale imaging data, showing consistent associations with a favourable metabolic profile, but novel associations with cardiac measures.
d) These findings highlight the limitations of using IGFBP-1 and IGFBP-2 as biomarkers in cardiometabolic disease.

## 3. Introduction

The rates of diabetes mellitus are on the rise, with prevalence estimated to rise by over 12.2% in 2045 (up to 800 million people)(1). Over 95% of diabetes cases are attributed to type 2 diabetes. It is well established that type 2 diabetes increases the risk of developing cardiovascular disease (CVD)(2), but despite reductions in CVD incidence with improved diabetes management, the incidence of CVD and mortality remains higher than in counterparts without diabetes(3). Identifying biomarkers underpinning type 2 diabetes and CVD is crucial for enhancing prevention and monitoring strategies.

Insulin-like growth factor binding proteins (IGFBPs) are a family of seven circulating proteins which regulate insulin-like growth factor (IGF) bioavailability, tissue localisation and metabolic actions(4). IGFBPs can also mediate IGF-independent effects, for example through amino acid sequences which can bind membrane receptors or allow localisation to the nucleus(4). By modulating IGF-mediated insulin sensitisation, IGFBPs have been implicated in metabolic homeostasis(5). Recently, low IGFBP-1 and IGFBP-2 levels have been identified as causal mediators of polygenic type 2 diabetes risk(6). Both IGFBP-1 and −2 are expressed by the liver and negatively regulated by insulin(7; 8). They are both similar in that they possess an Arg-Gly-Asp (RGD) domain at the C-terminus, which is not present in other IGFBPs, to mediate IGF-independent effects(5). Studies have convincingly established low circulating IGFBP-1 and −2 levels as predictors of the development of abnormal glucose tolerance, prediabetes and type 2 diabetes(9–13). Furthermore, both IGFBP-1/-2 have been positively associated with higher high-density lipoprotein (HDL) levels(14), and negatively associated with triglycerides (TG) and low-density lipoprotein (LDL)(12; 15-17), obesity(12; 18; 19) and blood pressure(20–22). This would imply that elevated levels of circulating IGFBP-1/-2 improve cardiometabolic profiles.

Previous studies demonstrate conflicting evidence with regards to IGFBP-1 and −2 in survival and CVD (see supplementary Table 1 for comprehensive study details). Heald, *et al.* identified that low IGFBP-1 levels are associated with a higher prevalence of macrovascular diseases (stroke, peripheral vascular disease, myocardial infarction (MI)) in type 2 diabetes participants(20). Laughlin, *et al.* demonstrated that lower baseline IGFBP-1 was associated with higher ischemic heart disease-related mortality risk(23). In addition, van den Beld, *et al.* demonstrated that high baseline IGFBP-2 levels predicted better survival from all-cause mortality, however when models were adjusted for insulin resistance, the association was reversed(24). Contrastingly, several studies have suggested that high levels of IGFBP-1/-2 predict an increased risk of CVD events and mortality.

Elevated levels of IGFBP-1 predict CVD-mortality(25) and incident heart failure (HF)(26). Similarly, IGFBP-2 is a marker for HF(27). Following MI, elevated IGFBP-2 levels predict major adverse cardiovascular events(28). In studies assessing both IGFBP-1 and IGFBP-2, high levels of both proteins are markers for all-cause mortality(29).

These studies were commonly in at-risk populations (e.g. with prior disease or over an average of 65 years old) and in relatively small sample sizes. UK Biobank (UKB) is a comprehensive resource containing detailed information for approximately 50,000 participants with quantified levels of IGFBP-1/-2 from the UKB plasma proteomics project. Participants between the ages of 37 and 73 years have been enrolled for at least 15 years and have a relatively low baseline prevalence of CVD. In addition, UKB is the world’s largest imaging study, with cardiac and abdominal MRI data available for approximately 5,000 participants within the plasma proteomics cohort. We aimed to utilise the UKB resource to identify associations of IGFBP-1 and IGFBP-2 with incident diabetes, CVD-related mortality and macrovascular disease incidence and integrate observations with novel imaging data available. Such data provide a particular novelty to this study, allowing powerful insights for IGFBP-1 and IGFBP-2 as putative biomarkers for identifying people at risk of cardiometabolic disease in the general population.

## 4. Methods

### The UK Biobank cohort

UKB is a large-scale prospective cohort study of 503,317 participants aged between 37 and 73 years recruited between 2006 and 2010. Assessments were performed across 22 centres in England, Scotland and Wales. UKB is an open-access resource containing information ranging from genetic data to detailed lifestyle questionnaires accessible to approved researchers(30). UKB is funded through UK government bodies and charities. Detailed information about study design can be found at https://www.ukbiobank.ac.uk/. UKB was granted initial ethical approval from the NHS Research Ethics Service (11/NW/0382). We conducted this research using application ID 547717. Participants provided written informed consent in line with the Declaration of Helsinki.

### Plasma proteomics

Plasma proteomic profiles for 54,219 participants were made available in a recent update to the UKB-Pharma Proteomics Project (UKB-PPP)(31). 2,923 proteins were measured using the Olink Explore 3072. Baseline expression profiles of IGFBP-1, IGFBP-2 and cardiac biomarkers Troponin I (TNNI3) and N-terminal prohormone of brain natriuretic peptide (NT-proBNP) are available. Levels are expressed as normalised protein expression (NPX) in log_2_ scale. For survival analyses, IGFBP-1 and IGFBP-2 were split into quartiles.

### Prevalent and incident diabetes mellitus

Participants completed a touch-screen questionnaire and a nurse-led verbal interview to collect sociodemographic information and medical history. The following codes were used to identify participants with prevalent diabetes mellitus: “1607: diabetic nephropathy”, “1468: diabetic neuropathy/ulcers”, “1220: diabetes”, “1222: type 1 diabetes”, “1223: type 2 diabetes”, “1276: diabetic eye disease” as previously described(32). Due to possible misclassification and only 1.5% of participants classified as having type 1 diabetes, we pooled both types to ensure inclusion of all cases. Therefore, we refer to diabetes mellitus throughout.

Dates of first reported diabetes mellitus were generated by mapping International Classification of Diseases 10^th^ Revision (ICD-10) codes to medical/self-reported records and death registers. The following codes were used: “130706: insulin-dependent diabetes mellitus”, “130708: non-insulin-dependent diabetes mellitus”, “130712: other specified diabetes mellitus” and “130714: unspecified diabetes mellitus”. Participants with first reports prior to initial assessment were removed from the analysis to calculate the time to incidence.

### Cardiovascular death and disease incidence

Mortality was identified using field codes “40000” and “40001” for date and underlying cause of death. Death caused by CVD was defined as previously described(32), using ICD-10 codes I00–I99 (excluding infection codes).

We selected to investigate myocardial infarction (MI, “42000”), ST-elevation MI (STEMI, “42002”), non-ST-elevation MI (NSTEMI, “42004”), stroke (“42006”), ischemic stroke (“42008”), heart failure (HF, “131354”) and peripheral vascular disease (PVD, “131386”) incidence. All except HF and PVD were algorithmically coded outcomes defined by the UKB outcome adjudication group. Participants with an occurrence date prior to the initial assessment visit were excluded to calculate time to incidence.

### Baseline clinical measurements

Baseline IGFBP-1 and IGFBP-2 expression levels were compared to physiological and blood biochemistry parameters which are associated with CVD or diabetes, including, but not limited to, BMI, systolic (sBP) and diastolic blood pressure (dBP), HbA1c, triglyceride (TG), C-reactive protein and low-density lipoprotein (LDL) (supplementary methods for field codes).

### Cardiac MRI

Baseline IGFBP-1 and IGFBP-2 were compared to available measures for all four cardiac chambers (supplementary methods for field codes). Left ventricular (LV) myocardial mass (MM), LV wall thickness (WT), stroke volume (SV), end systolic volume (ESV) and end diastolic volume (EDV) were indexed by body surface area (BSA, instance 2, “22427”). Cardiac contractility index (CCI) was calculated by dividing mean sBP (instance 2, “4080”) by LVESV indexed by BSA as previously described(33). For CCI, eight participants were excluded with a CCI above three standard deviations (SD) from the mean.

### Aortic dimensions and distensibility

Aortic maximum area and diameter were indexed to height and BSA respectively. Aortic distensibility (“24120” and “24123”) and maximum area (“24118” and “24121”) were obtained from the cardiac MRI analysis pipeline(34). Abdominal aorta horizontal and vertical diameter (“31090” and “31091) were obtained from data derived previously which excluded participants with conditions, such as diabetes and CVD(35). For ascending and descending aorta distensibility (∼ 4,000 participants) 53 and 62 outlying participants respectively were excluded. Abdominal aorta horizontal and vertical diameter contained data for 123 participants.

### Arterial stiffness index (ASI) and carotid intima-medial thickness (CIMT)

Baseline ASI at rest (“21021”, 17496 participants after excluding 52 outlying participants) was measured non-invasively with the PulseTrace PCA2 infrared device. CIMT was measured by ultrasound using a CardioHealth Station (Panasonic Biomedical Sales Europe BV, Leicestershire, UK), with a linear array transducer and a frequency of 5-13MHz. The mean CIMT (∼ 6,000 participants) from the right side at 120, 150 (“22671”, “22674”) and 210, 240 degrees on the left side (“22677”, “22680”) were calculated.

### Abdominal MRI

Baseline IGFBP-1 and −2 were compared to abdominal MRI measures of body composition (∼ 5,000 participants) derived from AMRA^®^ Medical AB (Linköping, Sweden)(36). We used visceral adipose tissue (VAT, “22407”), abdominal subcutaneous adipose tissue volume (ASAT, “22408”), thigh fat-free muscle volume (FFMV, “22409”), abdominal fat ratio (AFR, “22434”), muscle fat infiltration (MFI, “22435”), weight-muscle ratio (“22433”), average fat referenced (FR) liver proton density fat fraction (PDFF) (“24352”) and total abdominal adipose tissue index (TAATi, “22432”).

### Statistical analyses

Analyses were performed within the UKB RAP virtual platform (https://ukbiobank.dnanexus.com) using RStudio, R version 4.4.0 (2024-04-24). Analyses were performed with tidyverse v2.0.0 (22-02-2023) unless otherwise specified. Participants within the UKB-PPP dataset who were lost to follow-up were excluded (132 participants). Chi-squared tests with Bonferroni-corrected pairwise post-hoc tests and Kruskal-Wallis tests with Dunn’s post-hoc tests were completed for categorical or numerical data respectively. Adjusted linear regression models were performed for associations of IGFBP-1/-2 with cardiometabolic characteristics. Kaplan-Meier curves and adjusted Cox regression models were carried out using survminer v0.5.0 (30-10-2024) and survival v3.5.8 (14-02-2024). Analyses were adjusted for established confounders for CVD and diabetes, in addition to those thought to affect IGFBPs and IGFBP clearance (IGF-1, HbA1c and cystatin C).

Incident diabetes and CVD were censored on 31 May 2022. CVD-related and all-cause mortality were censored on 31 December 2023. Statistical significance was defined as *P* < 0.05.

## 5. Results

### Baseline Population Characteristics

In the UKB cohort, 52,881 participants with a mean age of 56.8 years (± 0.04 SEM) had plasma proteomic data available for analysis after exclusion of those lost to follow-up. In total 2,913 participants had a recorded diagnosis of diabetes mellitus at baseline. Participants were split into equal quartiles for baseline IGFBP-1 and IGFBP-2 protein expression (Table 1). Higher IGFBP-1 and −2 levels were associated with higher age, a lower proportion of male participants and a higher proportion of current smokers. However, participants had lower blood pressure, TG, HbA1c and BMI in the higher quartiles for both IGFBPs. The higher quartiles of both proteins had a lower prevalence of diabetes, but a higher prevalence of prior PVD. For IGFBP-1 there was no difference in prior stroke or HF. For IGFBP-2, there was higher prevalence of stroke and HF in Q4. There was no difference in prior MI for IGFBP-2 quartiles, but a lower prevalence of MI in Q4 IGFBP-1.

**Table 1.**
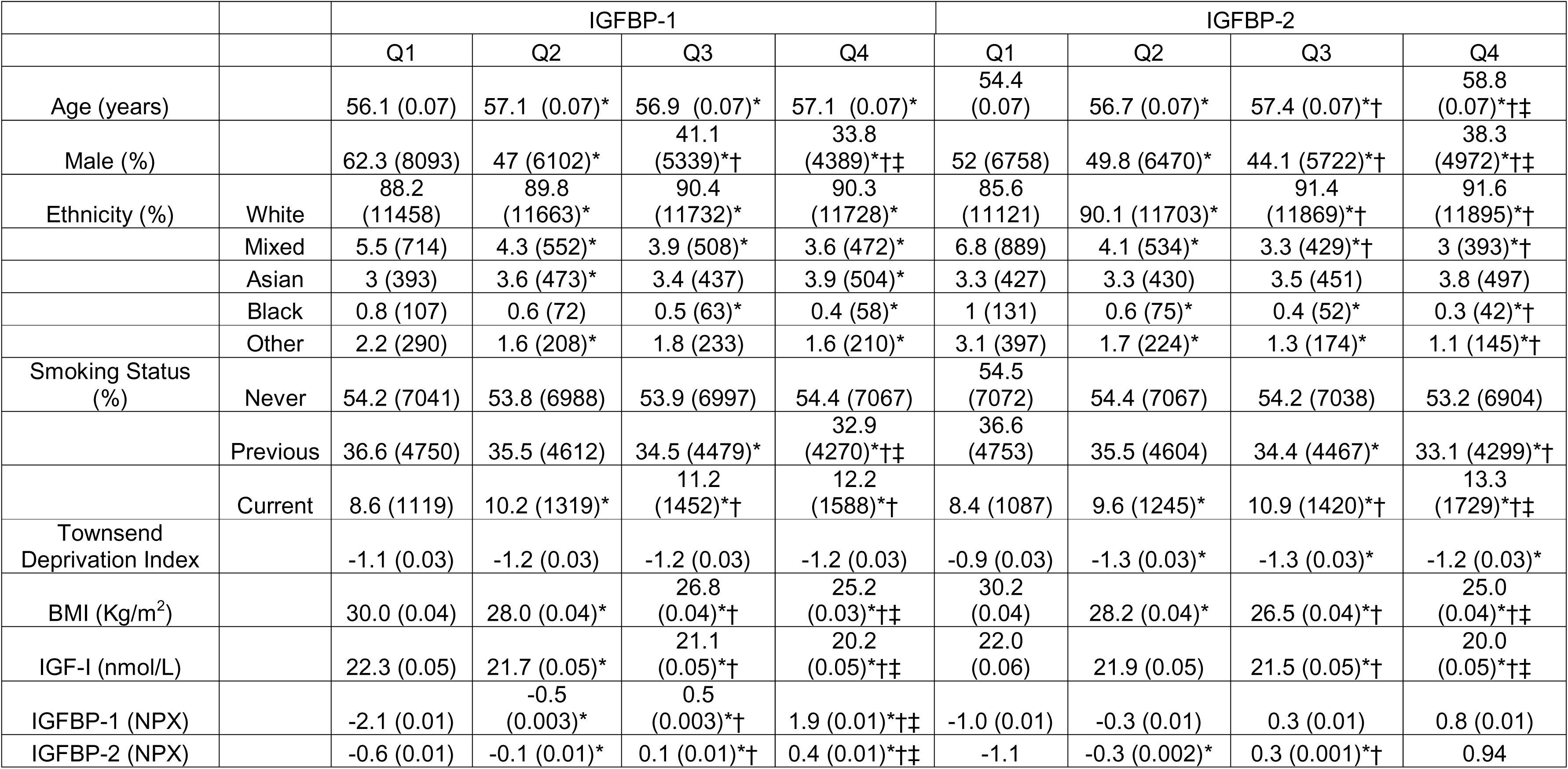
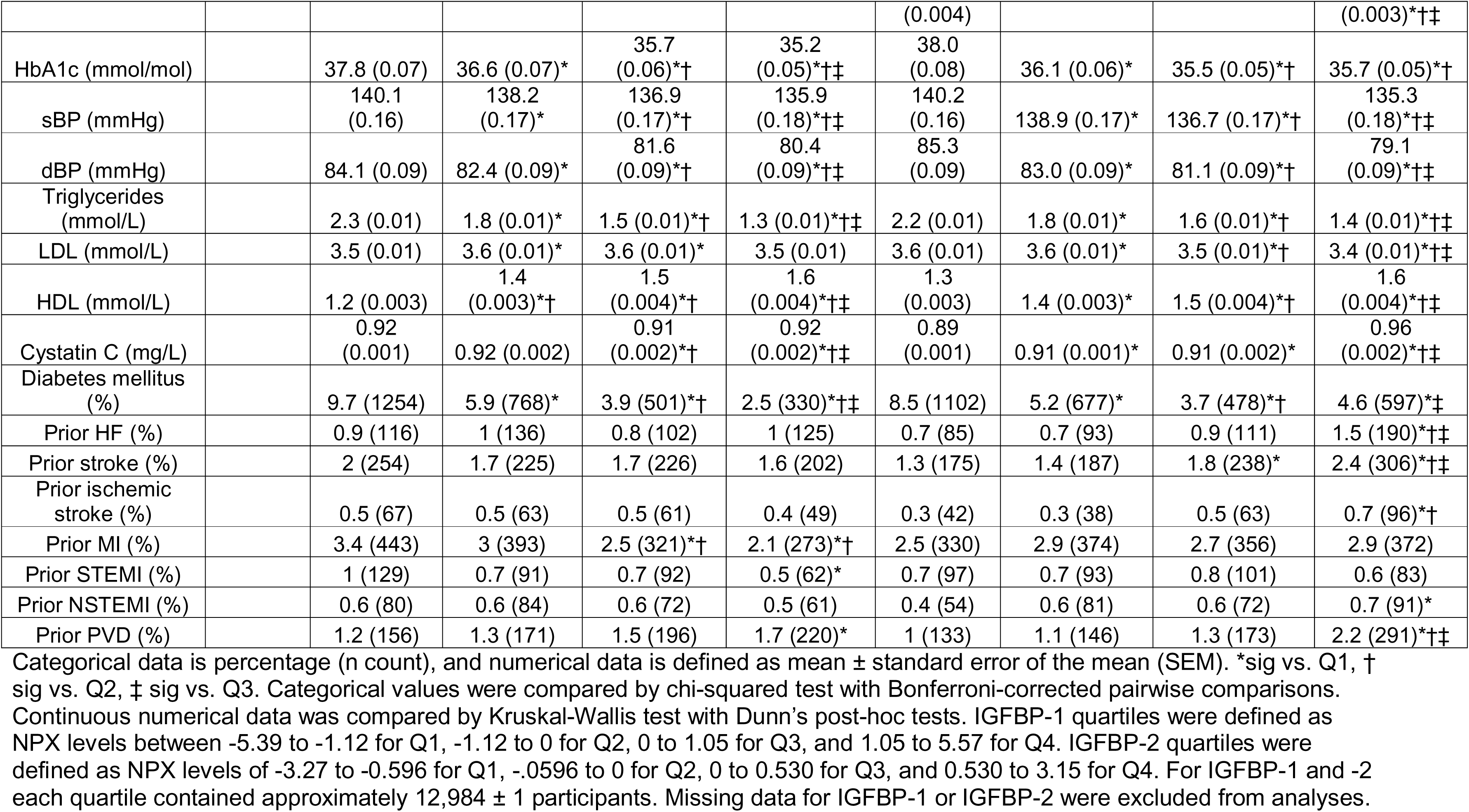
Baseline population characteristics of IGFBP-1 and IGFBP-2 quartiles.

### IGFBP-1 and −2 predict diabetes-free survival, but increased all-cause and cardiovascular-related mortality

During follow-up, 5,731 cohort participants died, of which 2,294 deaths were recorded as malignant neoplasms, followed by 1,204 CVD-related deaths (supplementary Table 2). After exclusion of participants with a diabetes diagnosis at baseline, 2,699 participants developed incident diabetes. Time-to-event was analysed for diabetes incidence, all-cause mortality and cardiovascular mortality to assess IGFBP-1/-2 as predictors of these outcomes. Both lower quartiles for IGFBP-1 and IGFBP-2 had an increased probability of diabetes mellitus diagnosis (Figure 1A-B). However, surprisingly, the inverse relationship was observed for all-cause (Figure 1C-D) and CVD-related mortality (Figure 1E-F). When CVD-caused mortality was split into subtypes, cardiovascular deaths were primarily caused by ischemic heart diseases (supplementary Figure 1).

**Figure 1.**
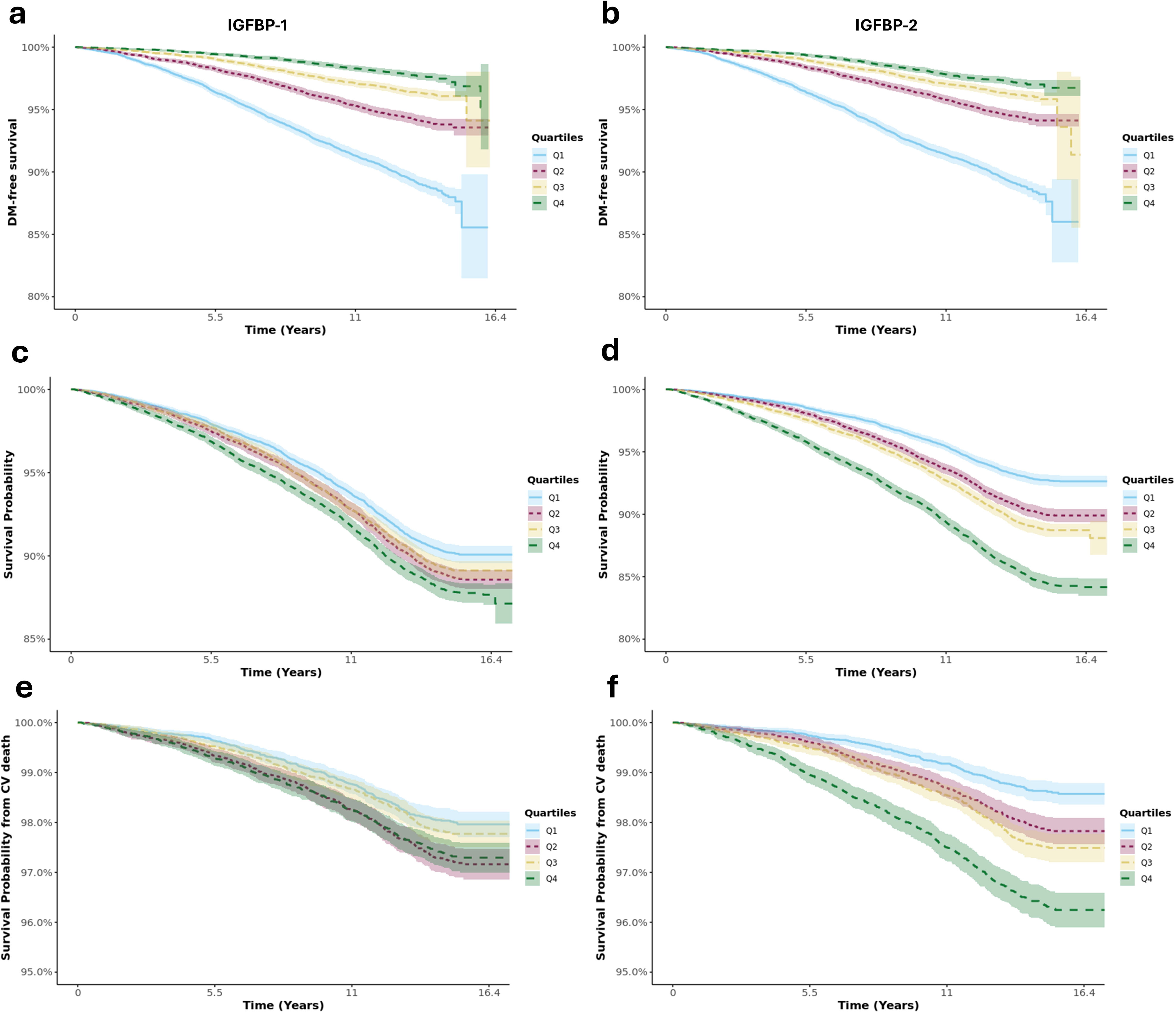
Unadjusted Kaplan-Meier curves representing survival probability from (a and b) diabetes mellitus **(DM)** incidence, (c and d) all-cause and (e and f) cardiovascular **(CV)** death. The population was split into quartiles of (a, c, e) **IGFBP-1** and (b, d, f) **IGFBP-2.**

Adjusted hazard ratios were obtained to assess risk associated with the different IGFBP quartiles (Table 2). Participants in Q4 had approximately 70% reduction in risk of developing diabetes (hazard ratio (HR) = 0.31 [95% CI 0.27 - 0.36] and HR = 0.32 [0.28 – 0.38] for IGFBP-1 and −2 respectively). Conversely, increasing quartiles in IGFBP-1 and −2 had a stepwise increase in risk of all-cause and cardiovascular mortality (HR = 1.81 [95% CI 1.47 – 2.24] and HR = 2.39 [95% CI 1.92 – 2.98] respectively).

**Table 2.**
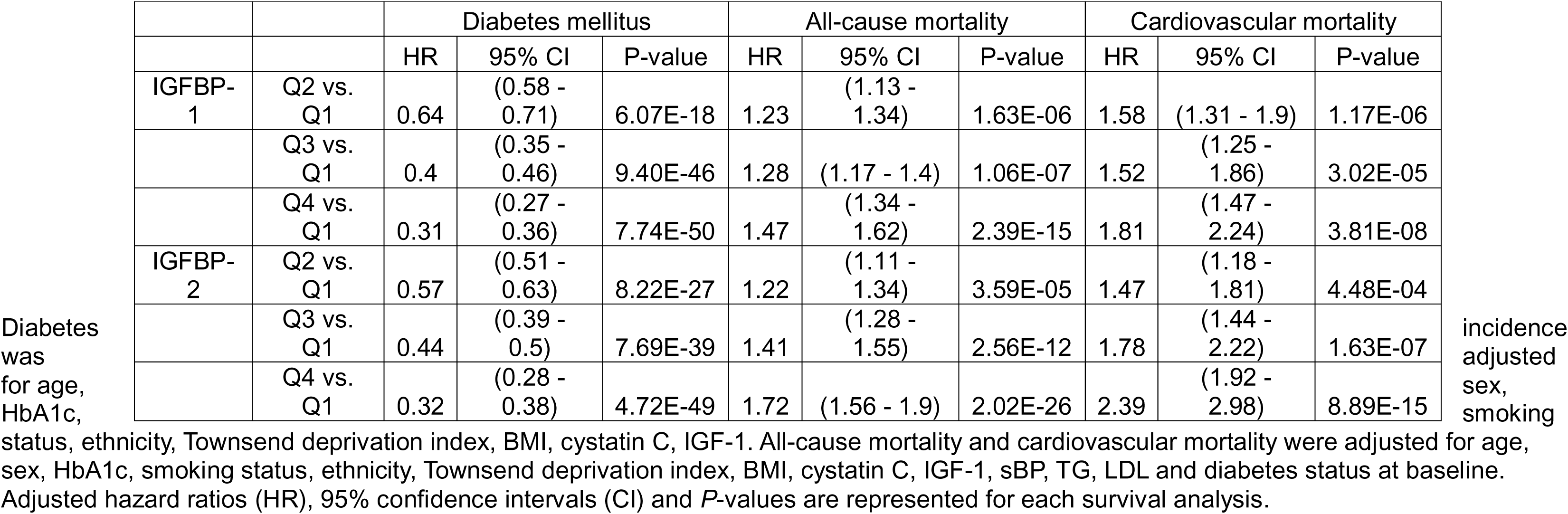
Results of multivariate Cox regression analyses of diabetes incidence, all-cause mortality and cardiovascular mortality.

We assessed whether the association of elevated baseline levels of IGFBP-1 and IGFBP-2 with an increased risk of mortality was not driven by participants without diabetes. Kaplan-Meier curves yielded persistent associations in the subset of participants with diabetes at baseline (Supplementary Figure 2). In adjusted analyses, elevated levels of IGFBP-1 and IGFBP-2 were associated with a substantial increased risk of CVD-related mortality in the diabetes subset (Table 3).

**Table 3.** Results of multivariate Cox regression analyses of cardiovascular mortality in the baseline diabetes cohort.

|  |  | HR | 95% CI | P-value |
| --- | --- | --- | --- | --- |
| IGFBP-1 | Q2 vs. Q1 | 1.75 | (1.09 - 2.80) | 1.95E-02 |
|  | Q3 vs. Q1 | 2.18 | (1.38 - 3.45) | 8.13E-04 |
|  | Q4 vs. Q1 | 1.99 | (1.22 - 3.23) | 5.45E-03 |
| IGFBP-2 | Q2 vs. Q1 | 1.22 | (0.72 - 2.07) | 4.56E-01 |
|  | Q3 vs. Q1 | 2.33 | (1.43 - 3.80) | 6.90E-04 |
|  | Q4 vs. Q1 | 2.81 | (1.70 - 4.67) | 5.45E-03 |

Cardiovascular mortality in the diabetes cohort was adjusted for age, sex, HbA1c, smoking status, ethnicity, Townsend deprivation index, BMI, cystatin C, IGF-1, sBP, TG and LDL. Adjusted hazard ratios (HR), 95% confidence intervals (CI) and *P*-values are represented for each survival analysis.

Additionally, sex-specific analyses for CVD-related mortality demonstrated that the associations persisted in the highest quartiles of IGFBP-1 and IGFBP-2, with similar hazard ratios for men and women (Table 4).

**Table 4.** Results of sex-specific multivariate Cox regression analyses of cardiovascular mortality in men and women.

|  |  | Men |  |  | Women |  |  |
| --- | --- | --- | --- | --- | --- | --- | --- |
|  |  | HR | 95% CI | P-value | HR | 95% CI | P-value |
| IGFBP-1 | Q2 vs. Q1 | 1.48 | (1.16 - 1.89) | 1.53E-03 | 1.92 | (1.39 - 2.65) | 8.23E-05 |
|  | Q3 vs. Q1 | 1.61 | (1.26 - 2.05) | 1.56E-04 | 1.27 | (0.87 - 1.86) | 2.14E-01 |
|  | Q4 vs. Q1 | 1.82 | (1.41 - 2.37) | 5.98E-06 | 2.12 | (1.47 - 3.05) | 6.21E-05 |
| IGFBP-2 | Q2 vs. Q1 | 1.66 | (1.25 - 2.19) | 3.78E-04 | 1.5 | (1.05 - 2.15) | 0.0271 |
|  | Q3 vs. Q1 | 1.85 | (1.39 - 2.46) | 2.34E-05 | 1.69 | (1.18 - 2.44) | 4.60E-03 |
|  | Q4 vs. Q1 | 2.77 | (2.08 - 3.68) | 2.34E-12 | 2.16 | (1.49 - 3.13) | 4.99E-05 |

Cardiovascular mortality in men and women in the UKB-PPP cohort was adjusted for age, HbA1c, smoking status, diabetes status at baseline, ethnicity, Townsend deprivation index, BMI, cystatin C, IGF-1, sBP, TG and LDL. Adjusted hazard ratios (HR), 95% confidence intervals (CI) and *P*-values are represented for each survival analysis.

### IGFBP-1 and −2 predict adverse CVD outcomes

Since higher concentrations of IGFBP-1 and −2 were associated with cardiovascular death, we investigated whether consistent trends were observed for incident macrovascular CVD in the whole cohort (Table 5). Participants in the highest quartile of IGFBP-1 and IGFBP-2 were more at risk of incident stroke, ischemic stroke, HF and PVD. However, only IGFBP-2 Q4 were 1.19 times more at risk of MI (p-value = 0.0343), which was borderline significant. When STEMI and NSTEMI were analysed separately this association was no longer observed (supplementary Table 3). Kaplan-Meier curves are available in supplementary figures 3, 4 and 5.

**Table 5.** Results of multivariate Cox regression analyses of CVD incidence.

|  |  | MI |  |  | Stroke |  |  | Ischemic Stroke |  |  | HF |  |  | PVD |  |  |
| --- | --- | --- | --- | --- | --- | --- | --- | --- | --- | --- | --- | --- | --- | --- | --- | --- |
|  |  | HR | 95% CI | P-value | HR | 95% CI | P-value | HR | 95% CI | P-value | HR | 95% CI | P-value | HR | 95% CI | P-value |
| IGFBP-1 | Q2 vs. Q1 | 1.15 | (1.01 - 1.31) | 0.0407 | 1.29 | (1.09 - 1.53) | 0.0035 | 1.27 | (1.06 - 1.53) | 0.00938 | 1.21 | (1.06 - 1.39) | 0.0047 | 1.02 | (0.83 - 1.25) | 0.846 |
|  | Q3 vs. Q1 | 1.02 | (0.88 - 1.18) | 0.807 | 1.16 | (0.97 - 1.39) | 0.114 | 1.22 | (1 - 1.48) | 0.0504 | 1.35 | (1.18 - 1.56) | 2.05E-05 | 1.41 | (1.15 - 1.72) | 0.000766 |
|  | Q4 vs. Q1 | 0.93 | (0.79 - 1.09) | 0.361 | 1.35 | (1.12 - 1.65) | 0.00221 | 1.32 | (1.07 - 1.63) | 0.00939 | 1.49 | (1.28 - 1.74) | 2.61E-07 | 1.53 | (1.23 - 1.89) | 0.000101 |
| IGFBP-2 | Q2 vs. Q1 | 1.15 | (1 - 1.33) | 0.0492 | 0.93 | (0.78 - 1.12) | 0.451 | 0.9 | (0.74 - 1.09) | 0.274 | 1.12 | (0.97 - 1.3) | 0.111 | 1.15 | (0.93 - 1.43) | 0.197 |
|  | Q3 vs. Q1 | 1.11 | (0.96 - 1.3) | 0.168 | 1.07 | (0.89 - 1.29) | 0.475 | 1.02 | (0.83 - 1.25) | 0.844 | 1.24 | (1.06 - 1.43) | 0.00535 | 1.44 | (1.16 - 1.79) | 0.000865 |
|  | Q4 vs. Q1 | 1.19 | (1.01 - 1.4) | 0.0343 | 1.26 | (1.04 - 1.53) | 0.0201 | 1.25 | (1.02 - 1.54) | 0.0326 | 1.56 | (1.34 - 1.82) | 1.69E-08 | 1.74 | (1.4 - 2.17) | 9.10E-07 |

All analyses were adjusted for age, sex, HbA1c, smoking status, ethnicity, Townsend deprivation index, BMI, cystatin C, IGF-1, sBP, TG, LDL and diabetes status at baseline. Adjusted hazard ratios (HR), 95% confidence intervals (CI) and *P*-values are represented for each survival analysis. MI = myocardial infarction, HF = heart failure, PVD = peripheral vascular disease.

### IGFBP-1 and IGFBP-2 are associated with a favourable metabolic phenotype, but possible alterations in cardiac phenotype

UKB is the largest whole-body imaging study, providing valuable insights into cardiac function to body fat composition. Using baseline measurements, blood biochemistry and MRI data, associations of IGFBP-1/-2 with cardiometabolic parameters were assessed. Both IGFBP-1 and IGFBP-2 were inversely associated with baseline waist/hip ratio and HbA1c (Table 6). In addition, both proteins were negatively associated with TG/HDL ratio, a surrogate marker of insulin resistance as previously described(37). IGFBP-1 and −2 were further compared to abdominal MRI measurements to assess fat composition. Higher levels of IGFBP-1 and IGFBP-2 were significantly associated with lower AFR, TAATi, VAT, ASAT and FR liver PDFF. The associations with body fat composition, were however modest. The negative associations per NPX unit increase of IGFBP-1 and IGFBP-2 were strongest for VAT and FR liver PDFF.

**Table 6.** Associations of IGFBP-1 and IGFBP-2 with metabolic phenotype markers.

|  | IGFBP-1 (NPX) |  | IGFBP-2 (NPX) |  |
| --- | --- | --- | --- | --- |
|  | Coefficient | P-value | Coefficient | P-value |
| HbA1c (mmol/mol) | -0.24 | < 2e-16 | -0.95 | < 2e-16 |
| TG/HDL ratio | -0.22 | < 2e-16 | -0.33 | < 2e-16 |
| waist/hip ratio | -0.01 | < 2e-16 | -0.02 | < 2e-16 |
| AFR | -0.01 | < 2e-16 | -0.02 | < 2e-16 |
| MFI (%) | -0.005 | NS | 0.03 | NS |
| TAATi (L/m<sup>2</sup>) | -0.07 | 5.17E-10 | -0.22 | < 2e-16 |
| VAT (L) | -0.15 | < 2e-16 | -0.48 | < 2e-16 |
| ASAT (L) | -0.06 | 0.004 | -0.14 | 0.001 |
| FFMV (L) | 0.05 | 0.005 | 0.05 | NS |
| FR liver PDFF mean (fraction) | -0.35 | 2.28E-16 | -1.45 | < 2e-16 |
| weight/FFMVratio | -0.02 | 0.040 | -0.01 | NS |

Multivariate linear regression models were performed to compare IGFBP-1 and IGFBP-2 levels to metabolic markers. HbA1c models were adjusted for age, sex, BMI, cystatin C, IGF-1, baseline diabetes status, smoking status, Townsend deprivation index and ethnicity. All other models were adjusted for age, sex, BMI, cystatin C, IGF-1, HbA1c, baseline diabetes status, baseline sBP, LDL, TG (except for TG/HDL ratio), smoking status, Townsend deprivation index and ethnicity. AFR = abdominal fat ratio, MFI = muscle fat infiltration, TAATi = total abdominal adipose tissue index, VAT = visceral adipose tissue, ASAT = abdominal subcutaneous adipose tissue, FFMV = fat-free muscle volume, FR liver PDFF = fat-referenced liver proton density fat fraction.

IGFBP-1 and IGFBP-2 were compared to surrogate markers of atherosclerotic disease from baseline measurements (Table 7). IGFBP-1 was not associated with blood pressure, while elevated levels of IGFBP-2 were negatively associated with blood pressure. Both proteins were negatively associated with ASI, but not CIMT. IGFBP-1 and IGFBP-2 were then compared to cardiac MRI measurements. There was no significant association of IGFBP-1 and −2 with ejection fraction (EF) in all four chambers, except for a borderline significant negative association of IGFBP-1 with left atrial EF. Both proteins showed a significant positive association with LVMMi. IGFBP-2 was negatively correlated with LVWTi, but this was not observed for IGFBP-1. IGFBP-1 and −2 were also negatively associated with CCI. Neither protein was associated with measurements of strain. However, both proteins were positively associated with SVi, ESVi and EDVi. In addition, elevated IGFBP-1 levels were positively associated with aortic area and abdominal aortic vertical diameter. IGFBP-2 was positively associated with aortic distensibility and aortic maximum area. Further to this, both proteins had a weak positive association with cardiac-specific Troponin I (marker of cardiomyocyte injury and MI) and a stronger positive association with NT-proBNP (cardiac wall stress, HF marker). IGFBP-1 was not associated with C-reactive protein (inflammatory marker of CVD risk), but IGFBP-2 demonstrated a negative association.

**Table 7.** Associations of IGFBP-1 and IGFBP-2 with cardiovascular phenotype markers.

|  | IGFBP-1 (NPX) |  | IGFBP-2 (NPX) |  |
| --- | --- | --- | --- | --- |
|  | Coefficient | P-value | Coefficient | P-value |
| sBP (mmHg) | 0.07 | NS | -2.35 | < 2e-16 |
| dBp (mmHg) | 0.06 | NS | -1.69 | < 2e-16 |
| ASI | -0.13 | 2.97E-14 | -0.30 | 3.38E-16 |
| CIMT ($\mu\text{m}$) | -0.80 | NS | 2.66 | NS |
| NT-proBNP (NPX) | 0.12 | < 2e-16 | 0.39 | < 2e-16 |
| C-reactive protein (mg/L) | 0.002 | NS | -0.17 | 5.26E-08 |
| Troponin I (NPX) | 0.01 | 3.53E-04 | 0.02 | 0.001 |
| **left ventricular chamber** |  |  |  |  |
| EF (%) | -0.05 | NS | -0.02 | NS |
| MMi ($\text{g}/\text{m}^2$) | 0.23 | 0.002 | 0.72 | 9.52E-06 |
| WTi ($\text{mm}/\text{m}^2$) | -0.005 | NS | -0.03 | 2.76E-04 |
| CCI (sBP/LVESVi) | -0.09 | 3.04E-09 | -0.26 | 7.72E-16 |
| CS (%) | 4.7E-05 | NS | 0.03 | NS |
| RS (%) | 0.03 | NS | 0.12 | NS |
| LS (%) | 0.02 | NS | -0.01 | NS |
| ESVi ($\text{mL}/\text{m}^2$) | 0.58 | 4.32E-09 | 1.68 | 7.40E-16 |
| EDVi ($\text{mL}/\text{m}^2$) | 1.26 | 2.76E-13 | 3.81 | < 2e-16 |
| SVi ($\text{mL}/\text{m}^2$) | 0.59 | 3.28E-09 | 2.17 | < 2e-16 |
| **left atrial chamber** |  |  |  |  |
| EF (%) | -0.22 | 0.044 | -0.04 | NS |
| SVi ($\text{mL}/\text{m}^2$) | 0.21 | 0.002 | 0.92 | 2.11E-10 |
| **right ventricular chamber** |  |  |  |  |
| EF (%) | 0.01 | NS | -0.11 | NS |
| EDVi ($\text{mL}/\text{m}^2$) | 1.24 | 1.19E-14 | 3.82 | < 2e-16 |
| ESVi ($\text{mL}/\text{m}^2$) | 0.53 | 4.57E-08 | 1.79 | < 2e-16 |
| SVi ($\text{mL}/\text{m}^2$) | 0.71 | 2.81E-12 | 2.03 | < 2e-16 |
| **right atrial chamber** |  |  |  |  |
| EF (%) | -0.04 | NS | 0.09 | NS |
| SVi ($\text{mL}/\text{m}^2$) | 0.49 | 2.57E-09 | 1.29 | 8.10E-14 |
| **aorta** |  |  |  |  |
| Ascending aorta distensibility ($10^{-3}/\text{mmHg}$) | 0.01 | 0.28 | 0.05 | 0.02 |
| Descending aorta distensibility ($10^{-3}/\text{mmHg}$) | 0.02 | 0.07 | 0.07 | 2.99E-03 |
| Ascending aorta maximum area ($\text{mm}^2/\text{m}$) | 0.06 | 1.19E-07 | 0.11 | 2.36E-05 |
| Descending aorta maximum area ($\text{mm}^2/\text{m}$) | 0.02 | 1.05E-04 | 0.03 | 2.71E-03 |
| Indexed abdominal aorta<br>horizontal diameter<br>(mm/m<sup>2</sup>) | 0.19 | 0.06 | 0.03 | 0.89 |
| Indexed abdominal aorta<br>vertical diameter (mm/m<sup>2</sup>) | 0.22 | 0.02 | 0.16 | 0.47 |

Multivariate linear regression models were performed to compare IGFBP-1 and IGFBP-2 levels to cardiovascular phenotype markers. Models were adjusted for age, sex, BMI, cystatin C, IGF-1, HbA1c, baseline diabetes status, baseline sBP (except for association with CCI, sBP and dBP), LDL, TG, smoking status, Townsend deprivation index and ethnicity. For aortic diameter, models were adjusted for age, sex, BMI, cystatin C, IGF-1, HbA1c, baseline sBP, LDL, TG, Townsend deprivation index and ethnicity. EF = ejection fraction, MMi = indexed myocardial mass, WTi = indexed wall thickness, CCI = cardiac contractility index, CS = circumferential strain, RS = radial strain, LS = longitudinal strain, ESVi = indexed end systolic volume, EDVi = indexed end diastolic volume, SVi = indexed stroke volume.

## 6. Discussion

Elevated circulating levels of IGFBP-1 and IGFBP-2 have been previously reported as markers of lower risk of incident diabetes, however contradictory associations have been reported with regards to their association with CVD. In our study, higher circulating levels of IGFBP-1 and IGFBP-2 have been identified as paradoxical putative biomarkers of lower diabetes incidence, with higher risk of all-cause and CVD-related mortality in the UKB cohort. Higher baseline levels of IGFBP-1 and −2 also indicate an increased risk of HF, PVD and stroke incidence. Despite CVD-related mortality being primarily caused by ischemic heart disease, IGFBP-1 and −2 did not predict a significant increased risk of incident STEMI or NSTEMI. Higher IGFBP-1 and IGFBP-2 were associated with a favourable body fat composition and metabolic profile. This is the first study of this scale to integrate associations of IGFBP-1 and IGFBP-2 with abdominal and cardiac MRI measures in all four cardiac chambers. This cardiometabolic paradox may indicate the IGFBP-1 and IGFBP-2 are not suitable biomarkers for identifying populations at risk of both diabetes and subsequent risk of CVD.

IGF-1 has been established as a mediator of survival, with its relationship believed to be U-shaped, with both low and high levels predicting mortality(38). Less than 1% of IGF-1 is thought to be free in circulation(5). IGFBP-3 is the most abundant high affinity IGFBP, sequestering 90% of circulating IGF-1 and IGF-2(4).

Fine-tuning of IGF bioactivity is thought to be mediated by other binding proteins, including IGFBP-2, the second most abundant(5), and then IGFBP-1. It is possible that elevated levels of IGFBP-1 and −2 are sequestering IGF-1 and are therefore associated with reduced survival from all-cause and CVD-related mortality in the UKB cohort. The associations that we observe in the UKB cohort may be IGF-mediated, although we adjusted for IGF-1 in all the analyses.

Consistent with previous reports that low levels of IGFBP-1 and IGFBP-2 predict diabetes incidence and development of impaired glucose tolerance(9–13), we observed the same association with diabetes incidence. Lower IGFBP-1 and −2 are convincingly established as markers for adverse cardiometabolic risk profiles. Both higher levels of IGFBP-1 and IGFBP-2 have previously been associated with lower BMI and TG and positively associated with HDL-cholesterol and insulin sensitivity(10; 12; 13; 18; 39; 40). These studies are in line with our findings that both proteins negatively associate with HbA1c, BMI, waist/hip ratio and TG/HDL ratio.

Overall, low baseline IGFBP-1 and −2 levels are cross-sectionally markers for reduced insulin sensitivity, glucose intolerance and dyslipidaemia. The mechanisms of IGFBP-1 and −2 involvement in insulin sensitivity have been investigated at the cellular and molecular level. Recombinant human (h)IGFBP-1 has previously been demonstrated to enhance insulin sensitivity by increasing insulin-stimulated phosphorylation of Akt and glucose uptake in C2C12 skeletal muscle cells and promoting insulin secretion from islet β-cells(41). Whole-body IGFBP-2 overexpression in mice protects from obesity and age-related glucose/insulin intolerance(42). IGFBP-2 is thought to mediate its effects by impairing adipogenesis(42), enhancing GLUT4 translocation and insulin responsiveness of adipocytes(43).

The observation that both elevated IGFBP-1 and −2 were associated with lower total adiposity, VAT, ASAT, AFR and hepatic fat content (FR liver PDFF) further corroborated the assumption that both IGFBPs are associated with a favourable metabolic profile. Previously both proteins have been negatively associated with VAT and total percent fat(29). Furthermore, high circulating IGFBP-2 levels have been associated with reduced liver fat content in men and women(44). A limitation of our study is that IGFBP expression may be dependent on the ambient metabolic health of the individual at baseline, which may change over time and with more advanced disease.

Few studies have identified associations of IGFBP-1 and IGFBP-2 with cardiac function in the general population. We observed no association of baseline IGFBP-1 and −2 levels with EF in all four cardiac chambers, but there was a negative association with LV CCI. We did not note an association of either protein with LV measures of strain. This may indicate a low prevalence of advanced ventricular dysfunction in this relatively healthy cohort. This is the first study using cardiac MRI data identifying a positive association of both IGFBP-1 and IGFBP-2 with ESVi, EDVi and SVi in all four cardiac chambers. Aortic distensibility and maximum area were also higher with elevated IGFBP-2, and IGFBP-1 was positively associated with aortic area and diameter. These observations are not causal, however, both IGFBPs appear to be associated with possible cardiac and aortic enlargement. Previously, higher IGFBP-2 has been associated with reduced LVEF in participants with acute coronary syndrome(28), and HF(27). Furthermore, IGFBP-2 has been associated with LV remodelling in aortic stenosis. Carter, *et al.* observed in asymptomatic aortic stenosis patients that IGFBP-2 is positively associated with SVi(45), which is consistent with our observations. Baseline levels of both IGFBP-1 and −2 were positively associated NT-proBNP levels and troponin I, which leads us to speculate that IGFBP-1 and IGFBP-2 may be associated with early cardiac stress or injury. This association, however, does not translate to reduced LVEF.

Recently, the role of IGFBP-1 has been investigated *in vitro* and *in vivo* in the context of cardiomyocyte HIF-1α mediated apoptosis in ischemia(46). In mice, six hours following coronary artery ligation, shRNA-mediated knockdowns of IGFBP-1 resulted in reduced infarct size and resulted in longer-term improvement of LVEF(46). It is important to note that our observations do not infer cause-effect relationships. It is possible that IGFBP expression may alter in response to changes in cardiac function, rather than playing a causal role. In future, it will be insightful to assess changes in IGFBP levels and their association with repeat imaging data which may become available in UKB.

ASI and CIMT are markers of CVD risk and surrogate markers of atherosclerotic disease. A previous study by Leionen, *et al.* identified a negative relationship between IGFBP-1 and CIMT in type 2 diabetes participants(47). However, Petersson-Pablo, *et al.* previously assessed pulse wave velocity and CIMT in Swedish young adults. IGFBP-1 and IGFBP-2 were negatively associated with pulse-wave velocity, but not CIMT(48). We observed that neither protein is associated with CIMT, but both are negatively associated with ASI. The aortic MRI analyses revealed that the negative association observed for IGFBP-1 and −2 in relation to ASI and IGFBP-2 with blood pressure, might in part be explained by the association with higher aortic dimensions and distensibility. To our knowledge, this the first report of associations of IGFBP-1 and IGFBP-2 with aortic dimensions.

There are limitations to using the UKB cohort for studying IGFBPs. We could not adjust our analyses for insulin levels as these were not measured in the assessments. Since insulin is a negative regulator of IGFBP-1 and IGFBP-2 to differing degrees(8), circulating insulin levels may affect plasma IGFBP-1/-2 levels. Also, blood was drawn throughout the day, and participants were not fasted. Because of the large number of individuals recruited to UKB, timing of blood sampling in relation to nutritional intake is unlikely to have had a substantial effect on our findings. We have also adjusted for overt confounding factors, including but not limited to baseline diabetes status, circulating IGF-1, HbA1c, LDL and TG, to control for factors affecting IGFBP expression and biological effects. Despite adjustments, many of the significant effects persisted, indicating that IGFBP associations were independent of these factors. Furthermore, IGF-1 was measured, but the assay does not distinguish bound vs. unbound IGF-1/IGFBP complexes or post-translational modifications of IGFBPs. For future studies, these may be considered to further validate associations. UKB is a powerful resource for diabetes studies due to the high prevalence and incidence of diabetes, however diabetes is often misclassified in clinical records. We included both type 2 and type 1 diabetes as some participants had two dates of diagnosis with both non-insulin dependent and insulin-dependent diabetes. We believe that inconsistencies in the presence of diabetes will be minimal due to the large number of participants, the lower number of type 1 diabetes cases and the inclusion of diabetes comorbidity codes. The UKB cohort is generally healthier than the general UK population, which is a limitation to this study as findings may not be applicable when generalised for the whole population. Finally, although UKB is good for sex-bias due to a slightly higher number of female participants, it is not ethnically diverse (approximately 80-90% white). This may affect the application of the results of our study in different ethnic groups.

Baseline IGFBP-1 and IGFBP-2 are powerful predictors of protection from incident diabetes, but are associated with increased risk for CVD, all-cause and CVD-related mortality. This large-scale study helps to corroborate the paradoxical dissociation of metabolic and cardiovascular risk associated with both proteins in a population that may better reflect the general population compared to previous studies. We have integrated into our large-scale analysis both abdominal and cardiac MRI measures, revealing that both proteins are associated with a favourable metabolic phenotype, but elevated levels are associated with possible alterations in cardiac chamber volumes. IGFBP-1 and IGFBP-2 may have limited potential as biomarkers for diabetes and the associated risk of CVD with diabetes.

## Data Availability

All data produced in the present study are available upon reasonable request to the authors

https://www.ukbiobank.ac.uk/

## 7. Acknowledgements

This research has been conducted using the UK Biobank Resource under Application Number 547717. This work uses data provided by patients and collected by the NHS as part of their care and support. E.R.R-H and M.S.C-R were supported by 4-year British Heart Foundation PhD studentships (FS/4yPhD/F/22/34182 for E.R.R-H and FS/4yPhD/F/21/34153 for M.S.C-R). L.E.D.F was supported by Diabetes UK project grant (22/0006395). P.J.M was supported by a British Heart Foundation Intermediate Basic Science Research Fellowship (FS/18/38/33659) and BBSRC research grant (BB/V014358/1). This work was supported by the British Heart Foundation 4-year PhD program (FS/4yPhD/F/22/34182). Graphical abstract logo reproduced by kind permission of UK Biobank ©. S.B.W is the guarantor for this work and, as such, had full access to all the data in the study and takes responsibility for the integrity of the data and the accuracy of the data analysis.

## 8. Conflicts of interest

None declared.

## 9. Author Contributions

All authors contributed to the research conceptualisation and manuscript writing – review and editing. E.R.R-H performed methodology, formal analyses and writing – original draft. M.S.C-R contributed to methodology. R.M.C, L.E.D.F, K.J.S, P.J.M and S.B.W were involved in supervision and funding acquisition.

## 10. Data availability statement

UK Biobank data used in this research article (https://www.ukbiobank.ac.uk/) are available to other scientists after application.

## Notes

### Competing Interest Statement

The authors have declared no competing interest.

### Author Declarations

UK Biobank was granted initial ethical approval from the NHS Research Ethics Service (11/NW/0382). We conducted this research using application ID 547717.

## References

1. Sun H, Saeedi P, Karuranga S, Pinkepank M, Ogurtsova K, Duncan BB, Stein C, Basit A, Chan JCN, Mbanya JC, Pavkov ME, Ramachandaran A, Wild SH, James S, Herman WH, Zhang P, Bommer C, Kuo S, Boyko EJ, Magliano DJ: IDF Diabetes Atlas: Global, regional and country-level diabetes prevalence estimates for 2021 and projections for 2045. Diabetes Res Clin Pract 2022;183:109119

2. Arnold SV, Khunti K, Tang F, Chen H, Cid-Ruzafa J, Cooper A, Fenici P, Gomes MB, Hammar N, Ji L, Saraiva GL, Medina J, Nicolucci A, Ramirez L, Rathmann W, Shestakova MV, Shimomura I, Surmont F, Vora J, Watada H, Kosiborod M: Incidence rates and predictors of microvascular and macrovascular complications in patients with type 2 diabetes: Results from the longitudinal global discover study. American Heart Journal 2022;243:232–239

3. Rawshani A, Rawshani A, Franzén S, Eliasson B, Svensson AM, Miftaraj M, McGuire DK, Sattar N, Rosengren A, Gudbjörnsdottir S: Mortality and Cardiovascular Disease in Type 1 and Type 2 Diabetes. N Engl J Med 2017;376:1407–1418

4. Haywood NJ, Slater TA, Matthews CJ, Wheatcroft SB: The insulin like growth factor and binding protein family: Novel therapeutic targets in obesity & diabetes. Mol Metab 2019;19:86–96

5. Wheatcroft SB, Kearney MT: IGF-dependent and IGF-independent actions of IGF-binding protein-1 and −2: implications for metabolic homeostasis. Trends Endocrinol Metab 2009;20:153–162

6. Ritchie SC, Lambert SA, Arnold M, Teo SM, Lim S, Scepanovic P, Marten J, Zahid S, Chaffin M, Liu Y, Abraham G, Ouwehand WH, Roberts DJ, Watkins NA, Drew BG, Calkin AC, Di Angelantonio E, Soranzo N, Burgess S, Chapman M, Kathiresan S, Khera AV, Danesh J, Butterworth AS, Inouye M: Integrative analysis of the plasma proteome and polygenic risk of cardiometabolic diseases. Nature Metabolism 2021;3:1476–1483

7. Brismar K, Fernqvist-Forbes E, Wahren J, Hall K: Effect of insulin on the hepatic production of insulin-like growth factor-binding protein-1 (IGFBP-1), IGFBP-3, and IGF-I in insulin-dependent diabetes. The Journal of Clinical Endocrinology & Metabolism 1994;79:872–878

8. Clemmons DR, Snyder DK, Busby WH, Jr.: Variables Controlling the Secretion of Insulin-Like Growth Factor Binding Protein-2 in Normal Human Subjects*. The Journal of Clinical Endocrinology & Metabolism 1991;73:727–733

9. Brismar K, Hilding A, Ansurudeen I, Flyvbjerg A, Frystyk J, Östenson C-G: Adiponectin, IGFBP-1 and −2 are independent predictors in forecasting prediabetes and type 2 diabetes. Frontiers in Endocrinology 2023;Volume 13 - 2022

10. Lewitt MS, Hilding A, Ostenson CG, Efendic S, Brismar K, Hall K: Insulin-like growth factor-binding protein-1 in the prediction and development of type 2 diabetes in middle-aged Swedish men. Diabetologia 2008;51:1135–1145

11. Petersson U, Ostgren CJ, Brudin L, Brismar K, Nilsson PM: Low levels of insulin-like growth-factor-binding protein-1 (IGFBP-1) are prospectively associated with the incidence of type 2 diabetes and impaired glucose tolerance (IGT): the Söderåkra Cardiovascular Risk Factor Study. Diabetes Metab 2009;35:198–205

12. Wittenbecher C, Ouni M, Kuxhaus O, Jähnert M, Gottmann P, Teichmann A, Meidtner K, Kriebel J, Grallert H, Pischon T, Boeing H, Schulze MB, Schürmann A: Insulin-Like Growth Factor Binding Protein 2 (IGFBP-2) and the Risk of Developing Type 2 Diabetes. Diabetes 2018;68:188–197

13. Rajpathak SN, He M, Sun Q, Kaplan RC, Muzumdar R, Rohan TE, Gunter MJ, Pollak M, Kim M, Pessin JE, Beasley J, Wylie-Rosett J, Hu FB, Strickler HD: Insulin-like growth factor axis and risk of type 2 diabetes in women. Diabetes 2012;61:2248–2254

14. Narayanan RP, Fu B, Oliver RL, Siddals KW, Donn R, Hudson JE, White A, Laing I, Ollier WE, Heald AH, Gibson JM: Insulin-like growth factor-II and insulin-like growth factor binding protein-2 prospectively predict longitudinal elevation of HDL-cholesterol in type 2 diabetes. Ann Clin Biochem 2014;51:468–475

15. Harrela M, Koistinen R, Tuomilehto J, Nissinen A, Seppälä M: Low serum insulin-like growth factor-binding protein-1 is associated with an unfavourable cardiovascular risk profile in elderly men. Ann Med 2000;32:424–428

16. Rajpathak SN, McGinn AP, Strickler HD, Rohan TE, Pollak M, Cappola AR, Kuller L, Xue X, Newman AB, Strotmeyer ES, Psaty BM, Kaplan RC: Insulin-like growth factor-(IGF)-axis, inflammation, and glucose intolerance among older adults. Growth Horm IGF Res 2008;18:166–173

17. Carter S, Li Z, Lemieux I, Alméras N, Tremblay A, Bergeron J, Poirier P, Deshaies Y, Després JP, Picard F: Circulating IGFBP-2 levels are incrementally linked to correlates of the metabolic syndrome and independently associated with VLDL triglycerides. Atherosclerosis 2014;237:645–651

18. Gibson JM, Westwood M, Young RJ, White A: Reduced insulin-like growth factor binding protein-1 (IGFBP-1) levels correlate with increased cardiovascular risk in non-insulin dependent diabetes mellitus (NIDDM). The Journal of Clinical Endocrinology & Metabolism 1996;81:860–863

19. Yang J, Zhou W, Wu Y, Xu L, Wang Y, Xu Z, Yang Y: Circulating IGFBP-2 levels are inversely associated with the incidence of nonalcoholic fatty liver disease: A cohort study. J Int Med Res 2020;48:300060520935219

20. Heald AH, Siddals KW, Fraser W, Taylor W, Kaushal K, Morris J, Young RJ, White A, Gibson JM: Low circulating levels of insulin-like growth factor binding protein-1 (IGFBP-1) are closely associated with the presence of macrovascular disease and hypertension in type 2 diabetes. Diabetes 2002;51:2629–2636

21. Heald AH, Cruickshank JK, Riste LK, Cade JE, Anderson S, Greenhalgh A, Sampayo J, Taylor W, Fraser W, White A, Gibson JM: Close relation of fasting insulin-like growth factor binding protein-1 (IGFBP-1) with glucose tolerance and cardiovascular risk in two populations. Diabetologia 2001;44:333–339

22. de Kort SW, van Doorn J, van de Sande AG, Leunissen RW, Hokken-Koelega AC: Serum insulin-like growth factor-binding protein-2 levels and metabolic and cardiovascular risk factors in young adults and children born small for gestational age. J Clin Endocrinol Metab 2010;95:864–871

23. Laughlin GA, Barrett-Connor E, Criqui MH, Kritz-Silverstein D: The prospective association of serum insulin-like growth factor I (IGF-I) and IGF-binding protein-1 levels with all cause and cardiovascular disease mortality in older adults: the Rancho Bernardo Study. J Clin Endocrinol Metab 2004;89:114–120

24. van den Beld AW, Carlson OD, Doyle ME, Rizopoulos D, Ferrucci L, van der Lely AJ, Egan JM: IGFBP-2 and aging: a 20-year longitudinal study on IGFBP-2, IGF-I, BMI, insulin sensitivity and mortality in an aging population. European Journal of Endocrinology 2019;180:109–116

25. Harrela M, Qiao Q, Koistinen R, Tuomilehto J, Nissinen A, Seppälä M, Leinonen P: High serum insulin-like growth factor binding protein-1 is associated with increased cardiovascular mortality in elderly men. Horm Metab Res 2002;34:144–149

26. Janszky I, Hallqvist J, Ljung R, Hammar N: Insulin-like growth factor binding protein-1 is a long-term predictor of heart failure in survivors of a first acute myocardial infarction and population controls. Int J Cardiol 2010;138:50–55

27. Berry M, Galinier M, Delmas C, Fournier P, Desmoulin F, Turkieh A, Mischak H, Mullen W, Barutaut M, Eurlings LW, Van Wijk S, Brunner-La Rocca H-P, Caubere C, Butler J, Roncalli J, Evaristi MF, Cohen-Solal A, Seronde M-F, Escamilla R, Ferrières J, Koukoui F, Smih F, Rouet P: Proteomics analysis reveals IGFBP2 as a candidate diagnostic biomarker for heart failure. IJC Metabolic & Endocrine 2015;6:5–12

28. Wang W, Yu K, Zhao SY, Mo DG, Liu JH, Han LJ, Li T, Yao HC: The impact of circulating IGF-1 and IGFBP-2 on cardiovascular prognosis in patients with acute coronary syndrome. Front Cardiovasc Med 2023;10:1126093

29. Hu D, Pawlikowska L, Kanaya A, Hsueh WC, Colbert L, Newman AB, Satterfield S, Rosen C, Cummings SR, Harris TB, Ziv E: Serum insulin-like growth factor-1 binding proteins 1 and 2 and mortality in older adults: the Health, Aging, and Body Composition Study. J Am Geriatr Soc 2009;57:1213–1218

30. Sudlow C, Gallacher J, Allen N, Beral V, Burton P, Danesh J, Downey P, Elliott P, Green J, Landray M, Liu B, Matthews P, Ong G, Pell J, Silman A, Young A, Sprosen T, Peakman T, Collins R: UK biobank: an open access resource for identifying the causes of a wide range of complex diseases of middle and old age. PLoS Med 2015;12:e1001779

31. Sun BB, Chiou J, Traylor M, Benner C, Hsu Y-H, Richardson TG, Surendran P, Mahajan A, Robins C, Vasquez-Grinnell SG, Hou L, Kvikstad EM, Burren OS, Davitte J, Ferber KL, Gillies CE, Hedman ÅK, Hu S, Lin T, Mikkilineni R, Pendergrass RK, Pickering C, Prins B, Baird D, Chen C-Y, Ward LD, Deaton AM, Welsh S, Willis CM, Lehner N, Arnold M, Wörheide MA, Suhre K, Kastenmüller G, Sethi A, Cule M, Raj A, Kang HM, Burkitt-Gray L, Melamud E, Black MH, Fauman EB, Howson JMM, Kang HM, McCarthy MI, Nioi P, Petrovski S, Scott RA, Smith EN, Szalma S, Waterworth DM, Mitnaul LJ, Szustakowski JD, Gibson BW, Miller MR, Whelan CD, Alnylam Human G, AstraZeneca Genomics I, Biogen Biobank T, Bristol Myers S, Genentech Human G, GlaxoSmithKline Genomic S, Pfizer Integrative B, Population Analytics of Janssen Data S, Regeneron Genetics C: Plasma proteomic associations with genetics and health in the UK Biobank. Nature 2023;622:329–338

32. Brown OI, Drozd M, McGowan H, Giannoudi M, Conning-Rowland M, Gierula J, Straw S, Wheatcroft SB, Bridge K, Roberts LD, Levelt E, Ajjan R, Griffin KJ, Bailey MA, Kearney MT, Cubbon RM: Relationship Among Diabetes, Obesity, and Cardiovascular Disease Phenotypes: A UK Biobank Cohort Study. Diabetes Care 2023;46:1531–1540

33. Conning-Rowland MS, Giannoudi M, Drozd M, Brown OI, Yuldasheva NY, Cheng CW, Meakin PJ, Straw S, Gierula J, Ajjan RA, Kearney MT, Levelt E, Roberts LD, Griffin KJ, Cubbon RM: The diabetic myocardial transcriptome reveals Erbb3 and Hspa2 as a novel biomarkers of incident heart failure. Cardiovascular Research 2024;120:1898–1906

34. Bai W, Suzuki H, Huang J, Francis C, Wang S, Tarroni G, Guitton F, Aung N, Fung K, Petersen SE, Piechnik SK, Neubauer S, Evangelou E, Dehghan A, O’Regan DP, Wilkins MR, Guo Y, Matthews PM, Rueckert D: A population-based phenome-wide association study of cardiac and aortic structure and function. Nat Med 2020;26:1654–1662

35. Petersen SE, Aung N, Sanghvi MM, Zemrak F, Fung K, Paiva JM, Francis JM, Khanji MY, Lukaschuk E, Lee AM, Carapella V, Kim YJ, Leeson P, Piechnik SK, Neubauer S: Reference ranges for cardiac structure and function using cardiovascular magnetic resonance (CMR) in Caucasians from the UK Biobank population cohort. J Cardiovasc Magn Reson 2017;19:18

36. Borga M, Ahlgren A, Romu T, Widholm P, Dahlqvist Leinhard O, West J: Reproducibility and repeatability of MRI-based body composition analysis. Magnetic Resonance in Medicine 2020;84:3146–3156

37. Baneu P, Văcărescu C, Drăgan SR, Cirin L, Lazăr-Höcher AI, Cozgarea A, Faur-Grigori AA, Cri□an S, Gai□ă D, Luca CT, Cozma D: The Triglyceride/HDL Ratio as a Surrogate Biomarker for Insulin Resistance. Biomedicines 2024;12

38. Lin J, Yang L, Huang J, Liu Y, Lei X, Chen R, Xu B, Huang C, Dou W, Wei X, Liu D, Zhang P, Huang Y, Ma Z, Zhang H: Insulin-Like Growth Factor 1 and Risk of Cardiovascular Disease: Results From the UK Biobank Cohort Study. J Clin Endocrinol Metab 2023;108:e850–e860

39. Janssen JA, Stolk RP, Pols HA, Grobbee DE, Lamberts SW: Serum total IGF-I, free IGF-I, and IGFB-1 levels in an elderly population: relation to cardiovascular risk factors and disease. Arterioscler Thromb Vasc Biol 1998;18:277–282

40. Maddux BA, Chan A, Mandarino LJ, Goldfine ID, De Filippis EA: IGF-Binding Protein-1 Levels Are Related to Insulin-Mediated Glucose Disposal and Are a Potential Serum Marker of Insulin Resistance. Diabetes Care 2006;29:1535–1537

41. Haywood NJ, Cordell PA, Tang KY, Makova N, Yuldasheva NY, Imrie H, Viswambharan H, Bruns AF, Cubbon RM, Kearney MT, Wheatcroft SB: Insulin-Like Growth Factor Binding Protein 1 Could Improve Glucose Regulation and Insulin Sensitivity Through Its RGD Domain. Diabetes 2017;66:287–299

42. Wheatcroft SB, Kearney MT, Shah AM, Ezzat VA, Miell JR, Modo M, Williams SC, Cawthorn WP, Medina-Gomez G, Vidal-Puig A, Sethi JK, Crossey PA: IGF-binding protein-2 protects against the development of obesity and insulin resistance. Diabetes 2007;56:285–294

43. Assefa B, Mahmoud AM, Pfeiffer AFH, Birkenfeld AL, Spranger J, Arafat AM: Insulin-Like Growth Factor (IGF) Binding Protein-2, Independently of IGF-1, Induces GLUT-4 Translocation and Glucose Uptake in 3T3-L1 Adipocytes. Oxid Med Cell Longev 2017;2017:3035184

44. Rauzier C, Chartrand DJ, Alméras N, Lemieux I, Larose E, Mathieu P, Pibarot P, Lamarche B, Rhéaume C, Poirier P, Després JP, Picard F: Associations of insulin-like growth factor binding protein-2 with metabolic profile and hepatic fat deposition in asymptomatic men and women. Am J Physiol Endocrinol Metab 2023;325:E99–e105

45. Carter S, Capoulade R, Arsenault M, Bédard É, Dumesnil JG, Mathieu P, Pibarot P, Picard F: Relationship Between Insulin-Like Growth Factor Binding Protein-2 and Left Ventricular Stroke Volume in Patients With Aortic Stenosis. Canadian Journal of Cardiology 2015;31:1447–1454

46. Tang X, Jiang H, Lin P, Zhang Z, Chen M, Zhang Y, Mo J, Zhu Y, Liu N, Chen X: Insulin-like growth factor binding protein-1 regulates HIF-1α degradation to inhibit apoptosis in hypoxic cardiomyocytes. Cell Death Discov 2021;7:242

47. Leinonen ES, Salonen JT, Salonen RM, Koistinen RA, Leinonen PJ, Sarna SS, Taskinen MR: Reduced IGFBP-1 is associated with thickening of the carotid wall in type 2 diabetes. Diabetes Care 2002;25:1807–1812

48. Pettersson-Pablo P, Nilsson TK, Breimer LH, Hurtig-Wennlöf A: IGFBP-1 and IGFBP-2 are associated with a decreased pulse-wave velocity in young, healthy adults. BMC Cardiovascular Disorders 2021;21:131

