## Supplemental Data for "Discordant associations of IGF-binding proteins 1 & 2 with diabetes and cardiovascular disease: insights from UK Biobank"

### Supplementary Methods

##### Baseline clinical measurements

Body mass index (BMI, “21001”), height (“50”), waist (“48”) and hip circumference (“49”) were measured by certified staff at the assessment centre. The mean of two measurements of systolic (sBP, “4080”) and diastolic blood pressure (dBP, “4079”) were acquired with the Omron 705 IT automated blood pressure monitor. HbA1c (“30750”) was measured in packed red blood cells by high-performance liquid chromatography (HPLC) analysis on a Bio-Rad VARIANT II Turbo. Serum IGF-1 (“30770”) was measured by chemiluminescent immunoassay analysis on a DiaSorin LIASON XL. Serum cystatin C (“30720”) was measured by latex enhanced immunoturbidimetric analysis on a Siemens ADVIA 1800. All the following analyses described were performed on a Beckman Coulter AU5800. Triglyceride (TG) (“30870”) levels were quantified by enzymatic assay. C-reactive protein (“30710”) was measured by high-sensitivity immunoturbidimetric analysis. High-density lipoprotein (HDL) (“30760”) was measured by enzyme immune-inhibition analysis and low-density lipoprotein (LDL) (“30780”) was measured by enzymatic protective selection analysis.

##### Cardiac MRI procedure and field codes

Starting in 2014, imaging studies were performed aiming to scan 100,000 participants. Cardiac MRI was performed on a clinical wide bore 1.5 Tesla scanner (MAGNETOM Aera, Syngo Platform VD13A, Siemens Healthcare, Erlangen, Germany) during approximately a 20-minute scan without contrast or pharmacological intervention (1). Available cardiac measures for all four heart chambers are listed in supplementary methods Table 1. For LVEF, LVSV, LVEDV and LVESV, the Siemens *syngo* InlineVF fully automated analysis was used (2), and all other measurements were calculated using automated machine-learning-based analysis pipelines (3).

Supplementary methods Table 1. Cardiac MRI measures used in this study for all four heart chambers, with their field codes and number of participants available in the UKB-PPP cohort after exclusion of participants lost to follow-up. Number of outlying participants excluded from the analysis is recorded in the right column.

| **Left Ventricular (LV) Measures** | **Field Code** | **Number of Participants** | **Outliers excluded due to > 3 SD from the mean** |
| --- | --- | --- | --- |
| LV ejection fraction (LVEF) | 22420 | 5202 |  |
| LV myocardial mass (LVMM) | 24105 | 5275 |  |
| LV wall thickness (LVWT) | 24140 | 5271 |  |
| LV end diastolic volume (LVEDV) | 22421 | 5202 | 5 participants excluded for LVEDVi |
| LV end systolic volume (LVESV) | 22422 | 5202 | 15 participants excluded for LVESVi |
| LV stroke volume (LVSV) | 22423 | 5202 | 1 participant excluded for LVSVi |
| LV circumferential strain (LVCS) | 24157 | 5266 |  |
| LV radial strain (LVRS) | 24174 | 5266 |  |
| LV longitudinal strain (LVLS) | 24181 | 5136 |  |
| **Left Atrial (LA) Measures** |  |  |  |
| LA stroke volume (LASV) | 24112 | 5213 |  |
| LA ejection fraction (LAEF) | 24113 | 5213 |  |
| **Right ventricular (RV) Measures** |  |  |  |
| RV ejection fraction (RVEF) | 24109 | 5275 |  |
| RV stroke volume (RVSV) | 24108 | 5275 |  |
| RV end systolic volume (RVESV) | 24107 | 5275 |  |
| RV end diastolic volume (RVEDV) | 24106 | 5275 |  |
| **Right Atrial (RA) Measures** |  |  |  |
| RA ejection fraction (RAEF) | 24117 | 5213 |  |
| RA stroke volume (RASV) | 24116 | 5213 |  |

##### Abdominal MRI

Abdominal MRI from neck-to-knee was performed using the same 1.5T MAGNETOM Aera for cardiac MRI during approximately a 10-minute scan (4). Visceral adipose tissue (VAT, “22407”) was determined by measuring adipose tissue volume within the abdominal cavity, excluding adipose tissue outside the abdominal skeletal muscles and within the cavity and posterior of the spine and back muscles. Abdominal subcutaneous adipose tissue volume (ASAT, “22408”) was measured in the abdomen from the top of the femoral head to the top of the thoracic vertebrae T9. Fat-free muscle volume (FFMV, “22409”) was measured from the anterior thigh (quadriceps femoris, sartorius, and tensor fascia latae) and posterior thigh (gluteus muscles, iliacus, adductor muscles, and hamstring). Thigh fat-free muscle volume (FFMV) is defined as viable muscle tissue with voxels with fat fraction <50%. Abdominal fat ratio (AFR, “22434”) was calculated by dividing the sum of VAT and ASAT by the sum of VAT, ASAT and FFMV. Muscle fat infiltration (MFI, “22435”) was measured as the mean fat fraction in the FFMV of the anterior thigh muscle. Weight-muscle ratio (“22433”) was calculated by dividing body weight by FFMV. Average fat referenced (FR) liver proton density fat fraction (PDFF) (FR liver PDFF, “24352”) was measured in up to nine (and at least three) regions of interest in the liver, avoiding inhomogeneities, major vessels, and bile ducts (based on the IDEAL imaging protocol) (5). Total abdominal adipose tissue index (TAATi, “22432”) was defined as total abdominal fat (VAT + ASAT) divided by height squared.

##### References for supplementary methods

1)	Petersen SE, Matthews PM, Francis JM, Robson MD, Zemrak F, Boubertakh R, et al. UK Biobank's cardiovascular magnetic resonance protocol. J Cardiovasc Magn Reson. 2016;18:8.

2)	Suinesiaputra A, Sanghvi MM, Aung N, Paiva JM, Zemrak F, Fung K, et al. Fully automated left ventricular mass and volume MRI analysis in the UK Biobank population cohort: evaluation of initial results. The International Journal of Cardiovascular Imaging. 2018;34(2):281-91.

3)	Bai W, Suzuki H, Huang J, Francis C, Wang S, Tarroni G, et al. A population-based phenome-wide association study of cardiac and aortic structure and function. Nat Med. 2020;26(10):1654-62.

4)	Littlejohns TJ, Holliday J, Gibson LM, Garratt S, Oesingmann N, Alfaro-Almagro F, et al. The UK Biobank imaging enhancement of 100,000 participants: rationale, data collection, management and future directions. Nature Communications. 2020;11(1):2624.

5)	Wilman HR, Kelly M, Garratt S, Matthews PM, Milanesi M, Herlihy A, et al. Characterisation of liver fat in the UK Biobank cohort. PLoS One. 2017;12(2):e0172921.

### Supplementary Tables

Supplementary Table 1. Epidemiological studies assessing IGFBP-1 and/or IGFBP-2 as markers or predictors for metabolic syndrome, type 2 diabetes (T2D), cardiovascular disease (CVD) or mortality (excluding papers reporting associations in children, adolescents and gestational diabetes).

| Author (date) | Number of study participants | Population-type | Type of study |  |
| --- | --- | --- | --- | --- |
| Janssen (1998)(1) | 218 | healthy, 55-80 years old, men and women | cross-sectional | **low IGFBP-1 and/or IGFBP-2 are associated with insulin resistance, metabolic syndrome, T2D risk** |
| Mohamed-Ali (1999)(2) | 80 | non-insulin dependent diabetes mellitus, males and females | cross-sectional |  |
| Heald (2001)(3) | 272 | men and women, 25-74 years old, excl. known DM | cross-sectional |  |
| Kalme (2005)(4) | 335 | men, 70-89 years old | prospective cohort |  |
| Rajpathak (2008)(5) | 922 | men and women, aged 65 + | prospective cohort |  |
| Lewitt (2008)(6) | 355 | Swedish men, 35–56 years old, excl. DM | case-control prospective cohort |  |
| Petersson (2009)(7) | 664 | men and women, 40-59 years old, non-DM | prospective cohort |  |
| Yeap (2010)(8) | 3,980 | men, aged 70 + | cross-sectional |  |
| Lewitt (2010)(9) | 240 | women, 35–56 years old | case-control prospective cohort |  |
| de Kort (2010)(10) | 298 | young adults born small for gestational age (SGA), mean age 20.9 and SGA children, mean age 7.1 | cross-sectional |  |
| Gokulakrishnan (2012)(11) | 100 | 50 subjects with normal glucose tolerance (NGT) and 50 subjects with T2D | cross-sectional |  |
| Rajpathak (2012)(12) | 742 | women, 30–55 years old | nested case-control |  |
| Carter (2014)(13) | 379 | Caucasian men (no prior T2D), 20-65 years old | cross-sectional |  |
| Hjorteberg (2017)(14) | 99 | men and women with T2D recently diagnosed and matched controls, average age 58 years | case-control/cross-sectional |  |
| Wittenbecher (2018)(15) | 2,778 | men and women, 35-65 years old | nested case-control |  |
| van den Beld (2019)(16) | 539 | men and women around 55, 65 and 75 years old | longitudinal cohort |  |
| Zhang (2020)(17) | 840 | men and women, average age of 76.1 years old | prospective cohort |  |
| Yang (2020)(18) | 763 | men and women, hospital-based, average age 54 years old | cohort study |  |
| Brismar (2023)(19) | 450 | men and women, 35-56 years old | nested case-control (prospective study) |  |
| Gibson (1996)(20) | 74 | non-insulin dependent diabetes mellitus | cross-sectional | **low IGFBP-1 and/or IGFBP-2 are associated with CVD risk, CVD events, CVD-mortality or all-cause mortality** |
| Harrela (2000)(21) | 331 | men, 70-89 years old | cross-sectional |  |
| Heald (2002)(22) | 160 | T2D | cross-sectional |  |
| Laughlin (2004)(23) | 1,185 | 51-98 years old, men and women, mean age 74 years | prospective cohort |  |
| Narayanan (2014)(24) | 489 | Men and women, average 62.9 years old, with T2D | prospective cohort |  |
| Olszanecka (2017)(25) | 152 | perimenopausal women with essential HT, 40-60 years old, | case-control/cross-sectional |  |
| Lewitt (2008)(6) | 355 | Swedish men, 35–56 years old, excl. DM | case-control prospective cohort | **increasing IGFBP-1 and/or IGFBP-2 over time are associated with insulin resistance, T2D risk** |
| Lewitt (2010)(9) | 240 | women, 35–56 years old | case-control prospective cohort |  |
| Harrela (2002)(26) | 622 | men, 65-84 years old | prospective cohort study | **high IGFBP-1 and/or IGFBP-2 are associated with CVD risk, CVD events, CVD-mortality or all-cause mortality** |
| Wallander (2007)(27) | 575 | men and women with T2D and suspected acute MI | prospective randomised trial |  |
| Hassfeld (2007)(28) | 90 | men and women with idiopathic dilated cardiomyopathy | cohort study |  |
| Kaplan (2008)(29) | 1122 | men and women, aged 65 +, without prior CVD | prospective cohort study |  |
| Kaplan (2008)(30) | 566 incident congestive heart failure cases | men and women, aged 65 + | case-cohort |  |
| Hu (2009)(31) | 625 | men and women, aged 70 + and in good health at recruitment | prospective cohort study |  |
| Janszky (2010)(32) | 853 cases of MI | Swedish citizens, 45–70 years old, blood was taken 3 months after MI | case-control |  |
| Kaplan (2012)(33) | 997 | men and women, average 85.2 years old | longitudinal cohort study |  |
| van den Beld (2012)(34) | survivors (n=223) and non-survivors (n=180) | men, aged 73–94 years | prospective cohort study |  |
| Urbonaviciene (2014)(35) | 440 | men and women with symptomatic peripheral arterial disease, average age 65.7 years old | longitudinal cohort study |  |
| Berry (2015)(36) | 224 controls vs 112 HF cases, 10 vs 30 HF, 90 vs. 361 HF) | 3 different cohorts | cross-sectional |  |
| Kaplan (2017)(37) | 2268 | men and women, aged 65 +, free of DM and CVD, mean age 77.8 | longitudinal cohort study |  |
| Barutaut (2020)(38) | 870 | HF patients, men and women | longitudinal cohort study |  |
| Hjorteberg (2023)(39) | 859 treatment-naive and 558 metformin-treated persons enrolled in the Danish Centre for Strategic Research in T2D | men and women with recent onset T2D | longitudinal cohort study |  |
| Wang (2023)(40) | 277 | men and women with acute coronary syndrome who underwent coronary angiography, average 60.5 years old | prospective cohort study |  |
| van den Beld (2019)(16) | 539 | men and women around 55, 65 and 75 years old | longitudinal cohort study |  |
| Brankovic (2018)(41) | 263 | Patients with chronic heart failure, average age of 67 years | prospective cohort study |  |
| Li, et al. (2025)(42) | 465 | Patients with peripheral arterial disease, average age of 71 years | prospective cohort study |  |
| Kaplan (2007)(43) | 534 coronary events, 370 ischemic stroke events, and 1122 controls | men and women, aged 65+ | longitudinal cohort study | **IGFBP-1 levels did not predict risk of incident coronary events or stroke** |
| Faxen (2016)(44) | HFpEF 85, HFrEF 79, controls 136 | 3 different cohorts | case-cohort | **IGFBP-1 was increased in HFpEF and HFrEF, but not associated with outcomes in either HFpEF or HFrEF. IGFBP-1 correlated with severity of HF in both HFpEF and HFrEF** |
| Kalme (2005)(4) | 335 | men, 70-89 years old | prospective cohort | **low IGFBP-1 was associated with metabolic syndrome, but not DM. The lowest quartile of IGFBP-1 showed no excess risk of CVD mortality.** |
| Maggio (2013)(45) | 1197 | men and women, aged 65+ | longitudinal cohort study | **High IGFBP-1 was associated with all-cause mortality. IGF-1/IGFBP-1 ratio had no significant relationship with CVD-mortality.** |
| Ritsinger (2018)(46) | 180 | men and women without DM and glucose under 11 mmol/L, admitted for acute MI, median age 64 | cohort study | **IGFBP-1 was associated with all-cause mortality, but not CVD-caused death or CVD incidence.** |

##### References for Supplementary Table 1

1. Janssen JA, Stolk RP, Pols HA, Grobbee DE, Lamberts SW: Serum total IGF-I, free IGF-I, and IGFB-1 levels in an elderly population: relation to cardiovascular risk factors and disease. Arterioscler Thromb Vasc Biol 1998;18:277-282

2. Mohamed-Ali V, Pinkney JH, Panahloo A, Cwyfan-Hughes S, Holly JM, Yudkin JS: Insulin-like growth factor binding protein-1 in NIDDM: relationship with the insulin resistance syndrome. Clin Endocrinol (Oxf) 1999;50:221-228

3. Heald AH, Cruickshank JK, Riste LK, Cade JE, Anderson S, Greenhalgh A, Sampayo J, Taylor W, Fraser W, White A, Gibson JM: Close relation of fasting insulin-like growth factor binding protein-1 (IGFBP-1) with glucose tolerance and cardiovascular risk in two populations. Diabetologia 2001;44:333-339

4. Kalme T, Seppälä M, Qiao Q, Koistinen R, Nissinen A, Harrela M, Loukovaara M, Leinonen P, Tuomilehto J: Sex hormone-binding globulin and insulin-like growth factor-binding protein-1 as indicators of metabolic syndrome, cardiovascular risk, and mortality in elderly men. J Clin Endocrinol Metab 2005;90:1550-1556

5. Rajpathak SN, McGinn AP, Strickler HD, Rohan TE, Pollak M, Cappola AR, Kuller L, Xue X, Newman AB, Strotmeyer ES, Psaty BM, Kaplan RC: Insulin-like growth factor-(IGF)-axis, inflammation, and glucose intolerance among older adults. Growth Horm IGF Res 2008;18:166-173

6. Lewitt MS, Hilding A, Ostenson CG, Efendic S, Brismar K, Hall K: Insulin-like growth factor-binding protein-1 in the prediction and development of type 2 diabetes in middle-aged Swedish men. Diabetologia 2008;51:1135-1145

7. Petersson U, Ostgren CJ, Brudin L, Brismar K, Nilsson PM: Low levels of insulin-like growth-factor-binding protein-1 (IGFBP-1) are prospectively associated with the incidence of type 2 diabetes and impaired glucose tolerance (IGT): the Söderåkra Cardiovascular Risk Factor Study. Diabetes Metab 2009;35:198-205

8. Yeap BB, Chubb SA, Ho KK, Setoh JW, McCaul KA, Norman PE, Jamrozik K, Flicker L: IGF1 and its binding proteins 3 and 1 are differentially associated with metabolic syndrome in older men. Eur J Endocrinol 2010;162:249-257

9. Lewitt MS, Hilding A, Brismar K, Efendic S, Ostenson CG, Hall K: IGF-binding protein 1 and abdominal obesity in the development of type 2 diabetes in women. Eur J Endocrinol 2010;163:233-242

10. de Kort SW, van Doorn J, van de Sande AG, Leunissen RW, Hokken-Koelega AC: Serum insulin-like growth factor-binding protein-2 levels and metabolic and cardiovascular risk factors in young adults and children born small for gestational age. J Clin Endocrinol Metab 2010;95:864-871

11. Gokulakrishnan K, Velmurugan K, Ganesan S, Mohan V: Circulating levels of insulin-like growth factor binding protein-1 in relation to insulin resistance, type 2 diabetes mellitus, and metabolic syndrome (Chennai Urban Rural Epidemiology Study 118). Metabolism 2012;61:43-46

12. Rajpathak SN, He M, Sun Q, Kaplan RC, Muzumdar R, Rohan TE, Gunter MJ, Pollak M, Kim M, Pessin JE, Beasley J, Wylie-Rosett J, Hu FB, Strickler HD: Insulin-like growth factor axis and risk of type 2 diabetes in women. Diabetes 2012;61:2248-2254

13. Carter S, Li Z, Lemieux I, Alméras N, Tremblay A, Bergeron J, Poirier P, Deshaies Y, Després JP, Picard F: Circulating IGFBP-2 levels are incrementally linked to correlates of the metabolic syndrome and independently associated with VLDL triglycerides. Atherosclerosis 2014;237:645-651

14. Hjortebjerg R, Laugesen E, Høyem P, Oxvig C, Stausbøl-Grøn B, Knudsen ST, Kim WY, Poulsen PL, Hansen TK, Bjerre M, Frystyk J: The IGF system in patients with type 2 diabetes: associations with markers of cardiovascular target organ damage. European Journal of Endocrinology 2017;176:521-531

15. Wittenbecher C, Ouni M, Kuxhaus O, Jähnert M, Gottmann P, Teichmann A, Meidtner K, Kriebel J, Grallert H, Pischon T, Boeing H, Schulze MB, Schürmann A: Insulin-Like Growth Factor Binding Protein 2 (IGFBP-2) and the Risk of Developing Type 2 Diabetes. Diabetes 2018;68:188-197

16. van den Beld AW, Carlson OD, Doyle ME, Rizopoulos D, Ferrucci L, van der Lely AJ, Egan JM: IGFBP-2 and aging: a 20-year longitudinal study on IGFBP-2, IGF-I, BMI, insulin sensitivity and mortality in an aging population. European Journal of Endocrinology 2019;180:109-116

17. Zhang WB, Aleksic S, Gao T, Weiss EF, Demetriou E, Verghese J, Holtzer R, Barzilai N, Milman S: Insulin-like Growth Factor-1 and IGF Binding Proteins Predict All-Cause Mortality and Morbidity in Older Adults. Cells 2020;9

18. Yang J, Zhou W, Wu Y, Xu L, Wang Y, Xu Z, Yang Y: Circulating IGFBP-2 levels are inversely associated with the incidence of nonalcoholic fatty liver disease: A cohort study. J Int Med Res 2020;48:300060520935219

19. Brismar K, Hilding A, Ansurudeen I, Flyvbjerg A, Frystyk J, Östenson C-G: Adiponectin, IGFBP-1 and -2 are independent predictors in forecasting prediabetes and type 2 diabetes. Frontiers in Endocrinology 2023;Volume 13 - 2022

20. Gibson JM, Westwood M, Young RJ, White A: Reduced insulin-like growth factor binding protein-1 (IGFBP-1) levels correlate with increased cardiovascular risk in non-insulin dependent diabetes mellitus (NIDDM). The Journal of Clinical Endocrinology & Metabolism 1996;81:860-863

21. Harrela M, Koistinen R, Tuomilehto J, Nissinen A, Seppälä M: Low serum insulin-like growth factor-binding protein-1 is associated with an unfavourable cardiovascular risk profile in elderly men. Ann Med 2000;32:424-428

22. Heald AH, Siddals KW, Fraser W, Taylor W, Kaushal K, Morris J, Young RJ, White A, Gibson JM: Low circulating levels of insulin-like growth factor binding protein-1 (IGFBP-1) are closely associated with the presence of macrovascular disease and hypertension in type 2 diabetes. Diabetes 2002;51:2629-2636

23. Laughlin GA, Barrett-Connor E, Criqui MH, Kritz-Silverstein D: The prospective association of serum insulin-like growth factor I (IGF-I) and IGF-binding protein-1 levels with all cause and cardiovascular disease mortality in older adults: the Rancho Bernardo Study. J Clin Endocrinol Metab 2004;89:114-120

24. Narayanan RP, Fu B, Oliver RL, Siddals KW, Donn R, Hudson JE, White A, Laing I, Ollier WE, Heald AH, Gibson JM: Insulin-like growth factor-II and insulin-like growth factor binding protein-2 prospectively predict longitudinal elevation of HDL-cholesterol in type 2 diabetes. Ann Clin Biochem 2014;51:468-475

25. Olszanecka A, Dragan A, Kawecka-Jaszcz K, Fedak D, Czarnecka D: Relationships of insulin-like growth factor-1, its binding proteins, and cardiometabolic risk in hypertensive perimenopausal women. Metabolism 2017;69:96-106

26. Harrela M, Qiao Q, Koistinen R, Tuomilehto J, Nissinen A, Seppälä M, Leinonen P: High serum insulin-like growth factor binding protein-1 is associated with increased cardiovascular mortality in elderly men. Horm Metab Res 2002;34:144-149

27. Wallander M, Norhammar A, Malmberg K, Ohrvik J, Rydén L, Brismar K: IGF binding protein 1 predicts cardiovascular morbidity and mortality in patients with acute myocardial infarction and type 2 diabetes. Diabetes Care 2007;30:2343-2348

28. Hassfeld S, Eichhorn C, Stehr K, Naegele H, Geier C, Steeg M, Ranke MB, Oezcelik C, Osterziel KJ: Insulin-like growth factor-binding proteins 2 and 3 are independent predictors of a poor prognosis in patients with dilated cardiomyopathy. Heart 2007;93:359-360

29. Kaplan RC, McGinn AP, Pollak MN, Kuller L, Strickler HD, Rohan TE, Xue X, Kritchevsky SB, Newman AB, Psaty BM: Total insulinlike growth factor 1 and insulinlike growth factor binding protein levels, functional status, and mortality in older adults. J Am Geriatr Soc 2008;56:652-660

30. Kaplan RC, McGinn AP, Pollak MN, Kuller L, Strickler HD, Rohan TE, Cappola AR, Xue X, Psaty BM: High insulinlike growth factor binding protein 1 level predicts incident congestive heart failure in the elderly. Am Heart J 2008;155:1006-1012

31. Hu D, Pawlikowska L, Kanaya A, Hsueh WC, Colbert L, Newman AB, Satterfield S, Rosen C, Cummings SR, Harris TB, Ziv E: Serum insulin-like growth factor-1 binding proteins 1 and 2 and mortality in older adults: the Health, Aging, and Body Composition Study. J Am Geriatr Soc 2009;57:1213-1218

32. Janszky I, Hallqvist J, Ljung R, Hammar N: Insulin-like growth factor binding protein-1 is a long-term predictor of heart failure in survivors of a first acute myocardial infarction and population controls. Int J Cardiol 2010;138:50-55

33. Kaplan RC, Bùzková P, Cappola AR, Strickler HD, McGinn AP, Mercer LD, Arnold AM, Pollak MN, Newman AB: Decline in circulating insulin-like growth factors and mortality in older adults: cardiovascular health study all-stars study. J Clin Endocrinol Metab 2012;97:1970-1976

34. van den Beld AW, Blum WF, Brugts MP, Janssen JAMJL, Grobbee DE, Lamberts SWJ: High IGFBP2 levels are not only associated with a better metabolic risk profile but also with increased mortality in elderly men. European Journal of Endocrinology 2012;167:111-117

35. Urbonaviciene G, Frystyk J, Urbonavicius S, Lindholt JS: IGF-I and IGFBP2 in peripheral artery disease: results of a prospective study. Scand Cardiovasc J 2014;48:99-105

36. Berry M, Galinier M, Delmas C, Fournier P, Desmoulin F, Turkieh A, Mischak H, Mullen W, Barutaut M, Eurlings LW, Van Wijk S, Brunner-La Rocca H-P, Caubere C, Butler J, Roncalli J, Evaristi MF, Cohen-Solal A, Seronde M-F, Escamilla R, Ferrières J, Koukoui F, Smih F, Rouet P: Proteomics analysis reveals IGFBP2 as a candidate diagnostic biomarker for heart failure. IJC Metabolic & Endocrine 2015;6:5-12

37. Kaplan RC, Strizich G, Aneke-Nash C, Dominguez-Islas C, Bůžková P, Strickler H, Rohan T, Pollak M, Kuller L, Kizer JR, Cappola A, Li CI, Psaty BM, Newman A: Insulinlike Growth Factor Binding Protein-1 and Ghrelin Predict Health Outcomes Among Older Adults: Cardiovascular Health Study Cohort. The Journal of Clinical Endocrinology & Metabolism 2017;102:267-278

38. Barutaut M, Fournier P, Peacock WF, Evaristi MF, Caubère C, Turkieh A, Desmoulin F, Eurlings LWM, van Wijk S, Rocca H-PB-L, Butler J, Koukoui F, Dambrin C, Mazeres S, Le Page S, Delmas C, Galinier M, Jung C, Smih F, Rouet P: Insulin-like Growth Factor Binding Protein 2 predicts mortality risk in heart failure. International Journal of Cardiology 2020;300:245-251

39. Hjortebjerg R, Kristiansen MR, Brandslund I, Aa Olsen D, Stidsen JV, Nielsen JS, Frystyk J: Associations between insulin-like growth factor binding protein-2 and insulin sensitivity, metformin, and mortality in persons with T2D. Diabetes Res Clin Pract 2023;205:110977

40. Wang W, Yu K, Zhao SY, Mo DG, Liu JH, Han LJ, Li T, Yao HC: The impact of circulating IGF-1 and IGFBP-2 on cardiovascular prognosis in patients with acute coronary syndrome. Front Cardiovasc Med 2023;10:1126093

41. Brankovic M, Akkerhuis KM, Mouthaan H, Brugts JJ, Manintveld OC, van Ramshorst J, Germans T, Umans V, Boersma E, Kardys I: Cardiometabolic Biomarkers and Their Temporal Patterns Predict Poor Outcome in Chronic Heart Failure (Bio-SHiFT Study). The Journal of Clinical Endocrinology & Metabolism 2018;103:3954-3964

42. Li B, Shaikh F, Younes H, Abuhalimeh B, Zamzam A, Abdin R, Qadura M: The Prognostic Potential of Insulin-like Growth Factor-Binding Protein 1 for Cardiovascular Complications in Peripheral Artery Disease. In *Journal of Cardiovascular Development and Disease*, 2025

43. Kaplan RC, McGinn AP, Pollak MN, Kuller LH, Strickler HD, Rohan TE, Cappola AR, Xue X, Psaty BM: Association of total insulin-like growth factor-I, insulin-like growth factor binding protein-1 (IGFBP-1), and IGFBP-3 levels with incident coronary events and ischemic stroke. J Clin Endocrinol Metab 2007;92:1319-1325

44. Faxén UL, Hage C, Benson L, Zabarovskaja S, Andreasson A, Donal E, Daubert JC, Linde C, Brismar K, Lund LH: HFpEF and HFrEF Display Different Phenotypes as Assessed by IGF-1 and IGFBP-1. J Card Fail 2017;23:293-303

45. Maggio M, Cattabiani C, Lauretani F, Bandinelli S, De Vita F, Dall’Aglio E, Corsonello A, Lattanzio F, Paolisso G, Ferrucci L, Ceda GP: Insulin-Like Growth Factor-1 Bioactivity Plays a Prosurvival Role in Older Participants. The Journals of Gerontology: Series A 2013;68:1342-1350

46. Ritsinger V, Brismar K, Mellbin L, Näsman P, Rydén L, Söderberg S, Norhammar A: Elevated levels of insulin-like growth factor-binding protein 1 predict outcome after acute myocardial infarction: A long-term follow-up of the glucose tolerance in patients with acute myocardial infarction (GAMI) cohort. Diab Vasc Dis Res 2018;15:387-395

Supplementary Table 2. The six leading causes of death in the UKB-PPP cohort

| **Cause of death** | **Number of deaths** |
| --- | --- |
| Malignant neoplasms | 2294 |
| CVD-related death | 1204 |
| Diseases of the nervous system | 637 |
| Diseases of the respiratory system | 507 |
| Diseases of the digestive system | 228 |
| Mental and behavioural disorders | 186 |

Supplementary Table 3. Results of multivariate Cox regression analyses of STEMI and NSTEMI incidence. All analyses were adjusted for age, sex, HbA1c, smoking status, ethnicity, Townsend deprivation index, BMI, cystatin C, IGF-1, sBP, TG, LDL, diabetes mellitus status at baseline. Adjusted hazard ratios (HR), 95% confidence intervals (CI) and P-values are represented for each survival analysis.

|  |  | STEMI | | | NSTEMI | | |
| --- | --- | --- | --- | --- | --- | --- | --- |
|  |  | HR | 95% CI | P-value | HR | 95% CI | P-value |
| IGFBP-1 | Q2 vs. Q1 | 1.17 | (0.9 - 1.52) | 0.25 | 1.17 | (0.99 - 1.39) | 0.0731 |
|  | Q3 vs. Q1 | 0.96 | (0.72 - 1.29) | 0.791 | 1.01 | (0.84 - 1.22) | 0.915 |
|  | Q4 vs. Q1 | 0.89 | (0.64 - 1.22) | 0.469 | 1 | (0.82 - 1.24) | 0.967 |
| IGFBP-2 | Q2 vs. Q1 | 1.12 | (0.86 - 1.48) | 0.401 | 1.19 | (1 - 1.43) | 0.0562 |
|  | Q3 vs. Q1 | 0.95 | (0.7 - 1.28) | 0.725 | 1.01 | (0.83 - 1.23) | 0.934 |
|  | Q4 vs. Q1 | 0.95 | (0.68 - 1.32) | 0.762 | 1.21 | (0.98 - 1.49) | 0.0789 |

### Supplementary Figures

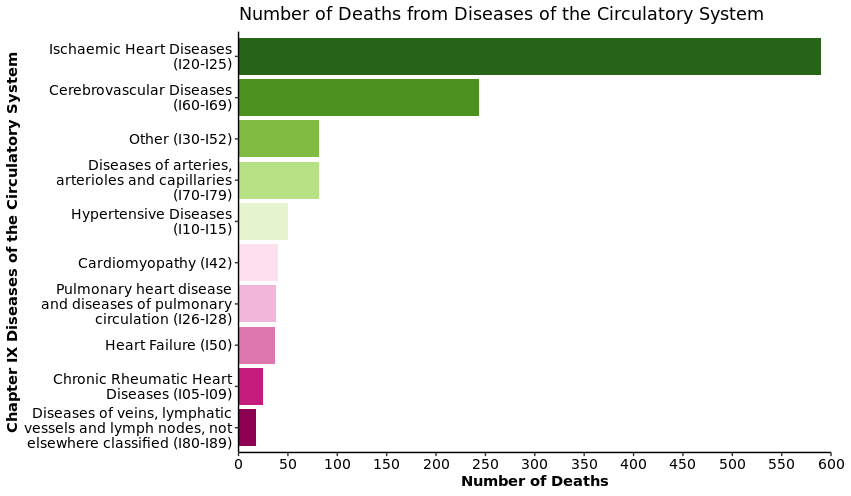

Supplementary Figure 1. Major causes of CVD-related deaths within Chapter IX of the ICD-10 death register codes (excluding deaths from infectious causes). CVDs were defined by the main ICD-10 grouping codes, except heart failure (I50) which was pulled out separately from Other CVDs (I30-I52) to identify participants who died of heart failure specifically. The total number of CVD-related deaths is 1,204 participants.

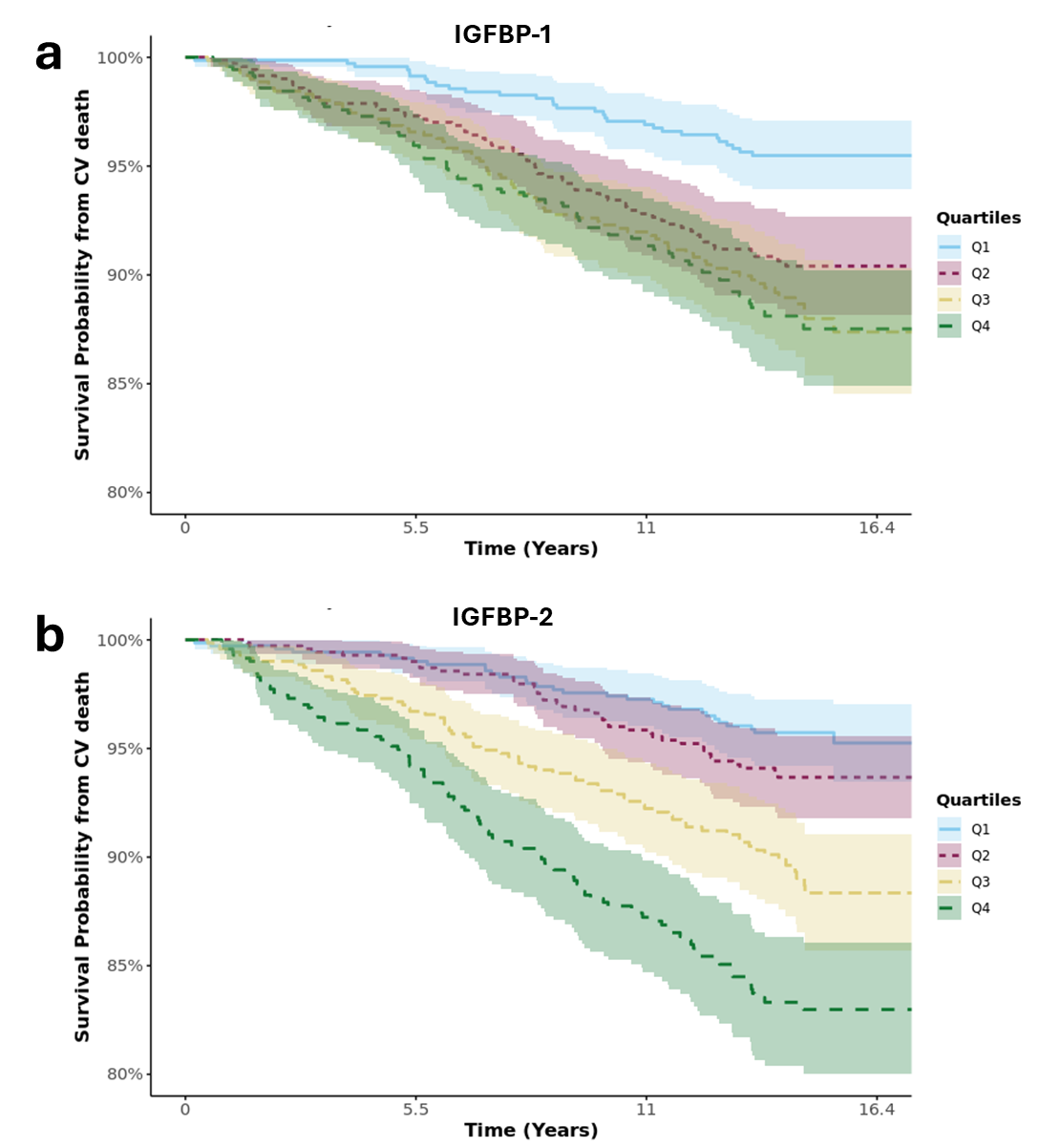

Supplementary Figure 2. Unadjusted Kaplan-Meier curves illustrating the survival from cardiovascular (CV)-related death in the subset of participants with diabetes at baseline. (A) Survival in IGFBP-1 quartiles. (B) Survival in IGFBP-2 quartiles.

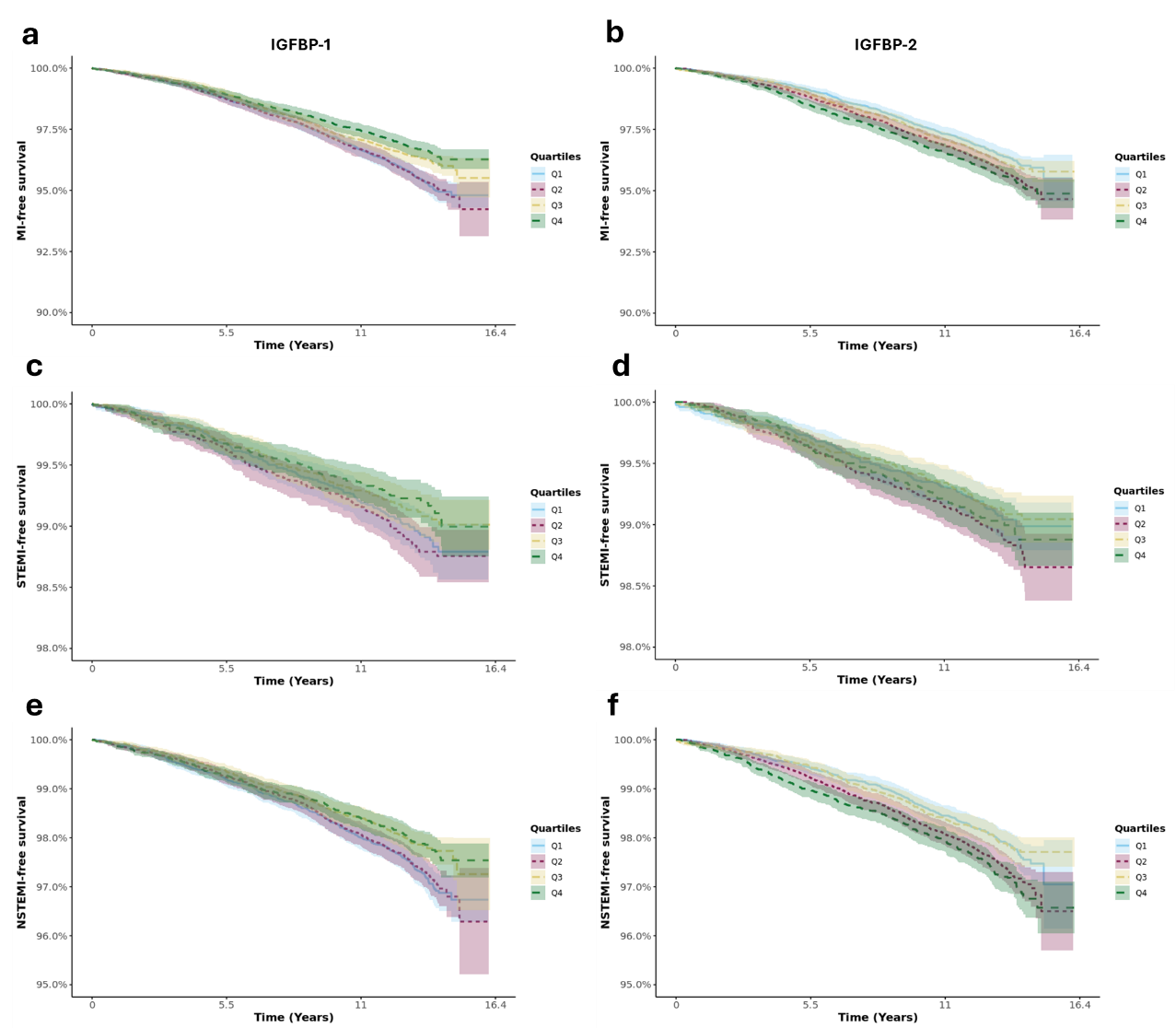

Supplementary Figure 3. Unadjusted Kaplan-Meier curves illustrating the survival from algorithmically coded myocardial infarction (A-B) MI, (C-D) STEMI, (E-F) NSTEMI. (A, C, E) Kaplan-Meier curves for IGFBP-1 and (B, D, F) Kaplan-Meier curves for IGFBP-2.

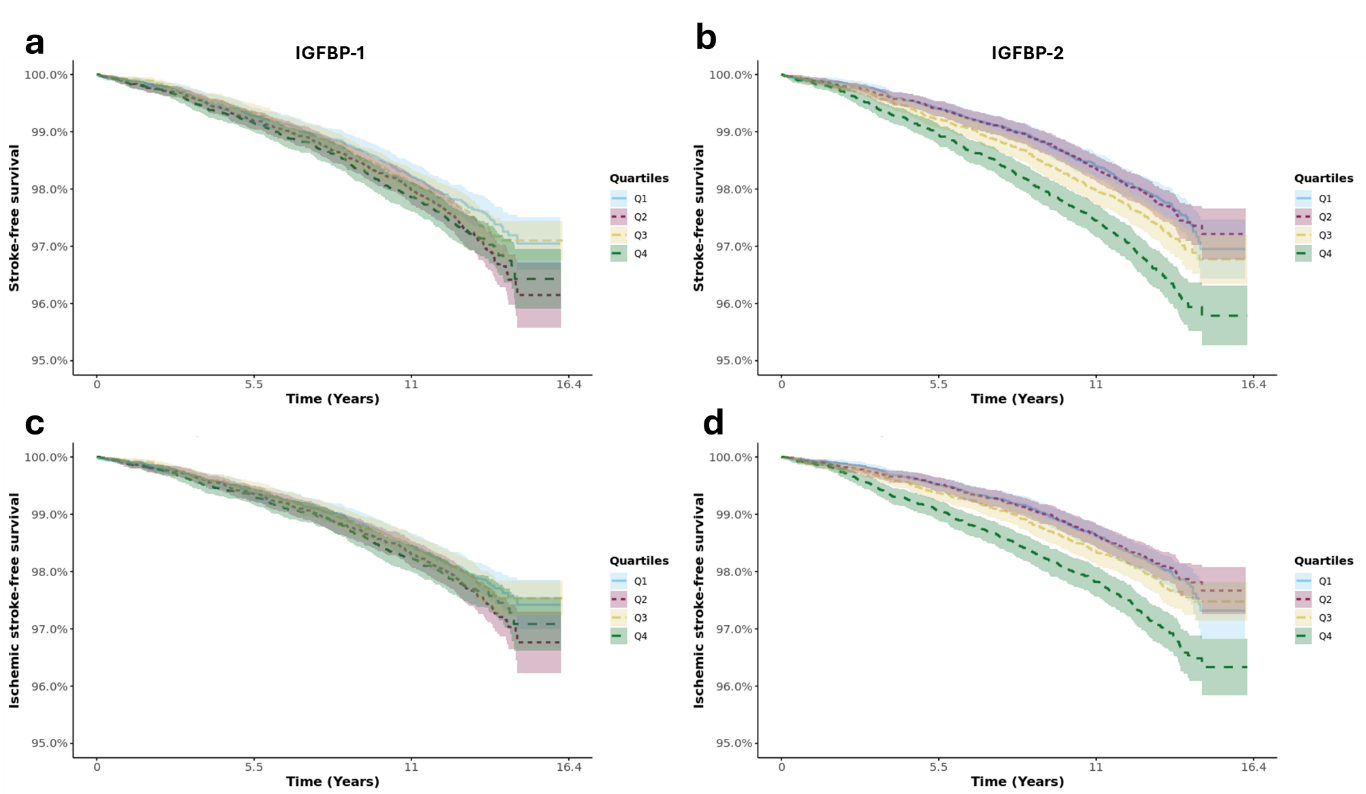

Supplementary Figure 4. Unadjusted Kaplan-Meier curves illustrating the survival from algorithmically coded (A-B) stroke and (C-D) ischemic stroke. (A, C) Kaplan-Meier curves for IGFBP-1 and (B, D) Kaplan-Meier curves for IGFBP-2.

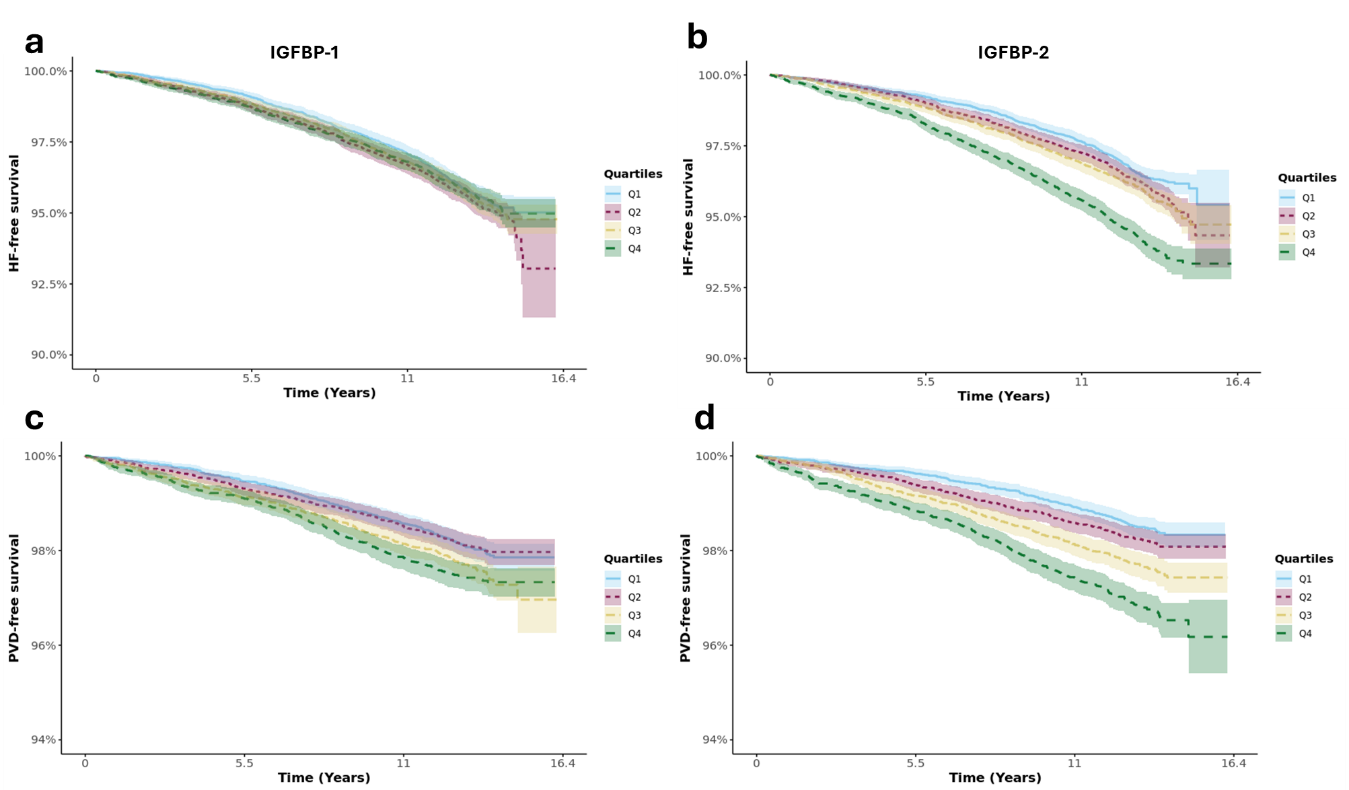

Supplementary Figure 5. Unadjusted Kaplan-Meier curves illustrating the survival from (A-B) heart failure (HF) and (C-D) peripheral vascular disease (PVD). (A, C) Kaplan-Meier curves for IGFBP-1 and (B, D) Kaplan-Meier curves for IGFBP-2.
